# Multi-ancestry analysis of *POLG* variants in Parkinson’s disease

**DOI:** 10.64898/2026.06.07.26354811

**Authors:** Yi Wen Tay, Inas Elsayed, Carlos F Hernandez, Dennis Yeow, Mikayla James, Pin-Jui Kung, Laurel Screven, Allison A. Dilliott, Roy N. Alcalay, Zih-Hua Fang, Ai Huey Tan, Global Parkinson’s Genetics Program (GP2), Carolyn M Sue, Lara M Lange, Maria Teresa Periñan

**Affiliations:** Department of Biomedical Science, Faculty of Medicine, University of Malaya, Kuala Lumpur, Malaysia; Faculty of Pharmacy, University of Gezira, Wad Medani, Sudan; Systems Biology Ireland, School of Medicine and Medical Science, University College Dublin, Dublin, Ireland; Universidad del Desarrollo, Centro de Genética y Genómica, Facultad de Medicina Clínica Alemana, Santiago 7610658, Chile; Neuroscience Research Australia, Randwick, New South Wales, Australia; Translational Neurogenomics Group, Garvan Institute of Medical Research, Sydney, NSW, Australia; Molecular Medicine Laboratory and Neurology Department, Concord Repatriation General Hospital, Concord, NSW, Australia; Faculty of Medicine and Health, University of Sydney, Camperdown, Australia; Center for Alzheimer’s and Related Dementias (CARD), National Institute on Aging and National Institute of Neurological Disorders and Stroke, National Institutes of Health, Bethesda, MD, USA; Division of Plastic Surgery, Department of Surgery, National Taiwan University Hospital, Taipei, Taiwan; The Global Parkinson’s Genetics Program (GP2), Bethesda, USA; Parkinson’s Foundation, NewYork, NY 10018, USA; Neurological Institute, Tel Aviv Sourasky Medical Center, Tel Aviv, Israel; Department of Neurology, Columbia University Irving Medical Center, NYC, NY 10032, USA; DataTecnica LLC, Washington, DC 20037, USA; Division of Neurology, Department of Medicine, and The Mah Pooi Soo & Tan Chin Nam Centre for Parkinson’s & Related Disorders, Faculty of Medicine, University of Malaya, Kuala Lumpur, Malaysia; Faculty of Medicine, University of New South Wales, Sydney, NSW, Australia; Department of Neurology, South Eastern Sydney Local Health District, Sydney, NSW, Australia; Department of Neurology, University Medical Center Hamburg-Eppendorf, Hamburg, Germany; Centre for Preventive Neurology, Wolfson Institute of Population Health, Queen Mary University of London, London, United Kingdom; Unidad de Trastornos del Movimiento, Servicio de Neurología y Neurofisiología Clínica, Instituto de Biomedicina de Sevilla, Hospital Universitario Virgen del Rocío/Consejo Superior de Investigaciones Científicas (CSIC)/Universidad de Sevilla, Seville, Spain

**Keywords:** *POLG*, Parkinson’s disease, genetics, multi-ancestry analysis

## Abstract

**Background:** Variants in the *polymerase gamma* (*POLG)* gene are associated with a wide range of mitochondrial disorders. Emerging evidence suggests a potential link between *POLG* variants and Parkinson’s disease (PD); yet, results remain inconclusive.

**Objective:** To investigate the spectrum and association of *POLG* genetic variation with PD across diverse ancestries.

**Methods:** We leveraged multi-ancestry data from the Global Parkinson’s Genetics Program (GP2), comprising genotyping data from 98,589 and short-read sequencing data from 36,022 individuals. We performed *POLG* rare pathogenic variant screen, CAG repeat analysis, gene-based rare variant and common variant association testing.

**Results:** Five PD patients carried homozygous or potentially compound heterozygous pathogenic or likely pathogenic *POLG* variants. Heterozygous pathogenic or likely pathogenic *POLG* variants were identified in 0.69% of PD patients and 0.51% of controls, without significant overall enrichment. The p.Thr251Ile-p.Pro587Leu pathogenic haplotype was similarly distributed between cases and controls. Although nominal associations were observed for individual variants and CAG repeat genotypes in specific ancestry groups, none survived correction for multiple testing. Gene-based rare variant analyses also revealed no significant associations with PD risk.

**Conclusions:** Rare pathogenic *POLG* variants were identified in a small proportion of individuals with PD, including five cases with potentially biallelic variants. However, we found no robust evidence supporting an association between *POLG* variation and PD susceptibility. These findings suggest that *POLG*-related mitochondrial dysfunction may contribute to PD in a small number of individuals, particularly when both copies of the gene are affected, although further clinical, genetic, and functional validation is needed.

**Plain language summary:** Mitochondria are the energy producers of our cells. The *POLG* gene helps keep them working. Changes in the POLG gene cause rare inherited diseases that affect muscles, nerves and the brain. Some people with these diseases also develop symptoms like those of Parkinson’s disease (PD). Earlier studies disagreed on whether POLG changes raise the risk of PD in people without these diseases.

We used genetic data from the Global Parkinson’s Genetics Program (GP2), which collects samples from people with and without PD worldwide. We used two types of data: a genetic chip test of about 98,600 people, and DNA sequencing (reading the DNA directly) of about 36,000 people. We looked for rare harmful POLG changes, common POLG changes, and differences in the length of a repeated stretch of DNA in the gene.

About 7 in 1,000 people with PD and 5 in 1,000 people without PD carried one harmful POLG change. A few other comparisons looked different at first, but none held up once we accounted for the many comparisons we made. Five people with PD carried two harmful POLG changes. For three of them, we could not confirm that the changes sat on different copies of the gene. We could not tell if these changes caused their PD.

We found no strong evidence that POLG changes raise the risk of PD in general. They may contribute to PD in a small number of cases. We had little clinical information about each person, so larger studies with detailed clinical information are needed.

## Introduction

The *polymerase gamma* (*POLG)* gene encodes the catalytic subunit of mitochondrial DNA (mtDNA) polymerase gamma, which is essential for mtDNA replication and maintenance.^1^ Genetic variants in *POLG* have been associated with mtDNA depletion or deletion, contributing to a broad spectrum of mitochondrial disorders, including progressive external ophthalmoplegia (PEO), Alpers syndrome, SANDO (sensory ataxia, neuropathy, deafness, and ophthalmoplegia), and other mitochondrial ataxia syndromes.^1,2^ To date, more than 300 pathogenic *POLG* variants have been reported, most of which predominantly associated with autosomal recessive disorders, whereas fewer exhibit autosomal dominant inheritance.

Beyond its established role in mitochondrial disorders, emerging evidence suggests a potential link between *POLG* and Parkinson’s disease (PD), possibly through impaired mtDNA maintenance leading to impaired mitochondrial function, increased oxidative stress, and neuronal loss in the substantia nigra.^3^ Postmortem brain analyses have revealed high levels of mtDNA deletions in dopaminergic neurons of the substantia nigra in both PD patients and aged individuals.^4–6^ In mouse models, intracerebral injection of damaged mtDNA induced PD-like symptoms that propagated in a prion-like manner.^6^ Moreover, patients with *POLG*-related mitochondrial disease exhibit marked neuronal cell loss in the substantia nigra, with α-synuclein pathology detected in some cases, further supporting a potential link between impaired mtDNA maintenance and neurodegeneration.^7^

Monoallelic and biallelic *POLG* variants have been reported in individuals with early- and late-onset PD and parkinsonism, sometimes accompanied by additional neurological manifestations, including external ophthalmoplegia and sensory neuropathy.^8–14^ In families affected by progressive external ophthalmoplegia, some *POLG* variants have also been found to co-segregate with parkinsonian symptoms.^3^ However, whether *POLG* variants contribute to idiopathic PD susceptibility remains unclear. Previous case-control studies investigating common *POLG* variants have generally found no significant enrichment in PD patients compared with controls.^15,16^ Associations between *POLG* exon 2 CAG repeat variation and PD have been reported in several populations, including Finnish, Norwegian, North American, Swedish, and Chinese.^15,17–21^ However, these findings have been inconsistent across studies,^22^ leaving the contribution of *POLG* genetic variation to PD risk unresolved.

In this study, we examined the frequency and genetic spectrum of rare disease-associated and common *POLG* variants, as well as exon 2 CAG repeat genotypes, in individuals with PD and other neurological phenotypes, and in controls across diverse ancestries. We addressed this by leveraging large-scale, multi-ancestry datasets from the Global Parkinson’s Genetics Program (GP2), including NeuroBooster Array (NBA) genotyping, short-read whole-genome sequencing (WGS), and clinical exome sequencing (CES).

## Methods

### Demographics

The demographics of the study cohort are detailed in Supplementary Table 1. We analyzed imputed NBA genotyping data from GP2 Data Release 11 (https://gp2.org/; DOI 10.5281/zenodo.17753486), comprising a total of 98,589 samples: 52,257 PD patients, 5,580 individuals with other phenotypes (including atypical parkinsonism and other neurodegenerative conditions), 39,470 healthy controls, and 1,282 unaffected family members of PD patients.

We further utilized the WGS dataset from GP2 Data Release 11, including 28,190 samples: 15,483 PD patients, 3,974 individuals with other phenotypes, 7,585 healthy controls, and 1,148 unaffected family members of PD patients. In addition, we incorporated CES data from 7,832 PD cases included in GP2 Data Release 11 generated by the Parkinson’s Foundation (PD GENEration) initiative, a large multi-center initiative based in North America.^23^ Individual cohort recruitment, sample collection, and data collection within GP2 have been described previously.^24–26^

All samples underwent genetic ancestry predictions using the GenoTools pipeline (https://github.com/GP2code/GenoTools), as previously described.^27^ Individuals were classified into eleven genetic ancestry groups: African Admixed (AAC), African (AFR), Ashkenazi Jews (AJ), Latinos and Indigenous People of the Americas (AMR), Complex Admixture History (CAH), Central Asian (CAS), East Asian (EAS), European (EUR), Finnish (FIN), Middle Eastern (MDE), and South Asian (SAS).

### Quality Control

For the NBA genotyping data, PLINK binary files were processed using the GenoTools pipeline, following standardized quality control (QC) procedures described previously.^27^ Briefly, samples were excluded if they failed any of the following criteria: call rate < 0.95, inconsistency between genetically inferred and reported sex, or excess heterozygosity (|F| > 0.15). After preliminary sample-level QC, variants with Hardy-Weinberg Equilibrium (HWE) ≤ 1 x 10^-5^ in control samples were removed. Variants showing non-random missingness by case-control status (P ≤ 1 x 10^-4^) or by haplotype (P ≤ 1 x 10^-4^) were also excluded.

For sequencing data, PLINK binary files were processed using the same GenoTools pipeline, following standardized QC procedures as described above. High-quality variants were retained using the following thresholds: call rate > 0.95, genotype quality >20, read depth >5, and heterozygous allele balance between 0.25 and 0.75.

### *POLG* rare pathogenic variant screening

*POLG* genomic coordinates were obtained from the Ensembl database (hg38: chr15: 89,305,198-89,334,861; https://www.ensembl.org).

For WGS and CES, *POLG* variants were extracted from variant call format (VCF) files using Bcftools (http://samtools.github.io/bcftools/howtos/publications.html) and annotated with ANNOVAR^28^ and Slivar (https://github.com/brentp/slivar). Slivar-specific parameters were used to filter variants by population allele frequencies, and rare variants with a gnomAD PopMax allele frequency <0.005 were retained.

Candidate variants were prioritized based on predicted deleteriousness and clinical pathogenicity classifications. Variants were retained if they met the following criteria: (i) located in coding or splice-affecting regions of *POLG*, including missense, predicted loss-of-function (stop-gain, stop-loss, frameshift, start-loss), or short insertion/deletion variants; (ii) Combined Annotation Dependent Depletion (CADD) score ≥12.37; and (iii) classification as pathogenic or likely pathogenic by at least two of three annotations resources: ClinVar^29^, Franklin^30^, and Varsome^31^, the latter two based on the consensus guideline of the American College of Medical Genetics and Genomics (ACMG).^32^

The previously described co-occurring *POLG* variants^33^ p.Thr251Ile and p.Pro587Leu were additionally assessed using raw genotyping data, with carriers identified through PLINK v2.0. For individuals with both genotyping and WGS data available, carrier status was cross-validated across platforms to ensure concordance.

### *POLG* CAG repeat analysis

The *POLG* exon 2 CAG repeat, which encodes a polymorphic polyglutamine (polyQ) tract, was analyzed using ExpansionHunter (EH) on GP2 R11 WGS CRAM files (GRCh38, chr15:89,333,590–89,333,629).^34^ The *POLG* polyQ tract consists of an uninterrupted CAG repeat followed by a CAA interruption and two additional CAG codons, represented as (CAG)n-CAA-CAG-CAG. To align with the previously used nomenclature for the uninterrupted CAG tract, three units were subtracted from the reported repeat length, accounting for the CAA interruption and two downstream CAG codons.^21^ Associations between *POLG* CAG repeat genotypes and PD risk were evaluated using three genotype comparisons: non-10/10 versus 10/10, non-11/11 versus 11/11, and non-10/11 versus 10/11. Logistic regression models were adjusted for age, sex, and the first five genetic principal components (PCs) except in the CAH, CAS, EUR, and MDE ancestry groups, which were adjusted only for sex and the first five PCs due to substantial missingness in age.

### Gene-based rare variant analysis

To evaluate the cumulative contribution of rare *POLG* variants to PD risk, we performed gene-based rare variant analyses leveraging the WGS data using the Combined Multivariate and Collapsing – Wald Test (CMC-Wald) and sequence kernel association test optimal unified SKAT (SKAT-O) methods as implemented in RVTESTS.^35^

Rare variants with MAF <1% were grouped into four masks: (i) potentially functional variants, including all except intronic variants; (ii) coding variants; (iii) loss-of-function variants (i.e., splicing, stop-gain, stop-loss, and frameshift), and (iv) variants with CADD >20. Gene-based association analyses were not performed in the AAC, CAH, CAS, FIN, and SAS ancestry groups due to the limited sample sizes (<1000) among PD patients and controls.

### Common variant association analysis

We further investigated the association between common *POLG* variants and PD risk by analyzing NBA genotyping imputed data. We used PLINK v2.0 for variant extraction and ANNOVAR^34^ for variant annotation; only variants with an imputation quality score (R²) ≥0.8 were kept for further analyses. Additional pruning was applied to remove rare variants with MAF < 1% in each ancestry group and variants with a minor allele count (MAC) <2. SNP-phenotype association analyses were conducted using a generalized linear model implemented in PLINK v2.0.^35^ Logistic regression models included sex, age, and 5 PCs as covariates to account for population stratification. Bonferroni correction was applied to adjust p-values for all *POLG* variants within each ancestry group.

## Results

### Potentially biallelic pathogenic or likely pathogenic *POLG* variant carriers

Five PD patients carried either homozygous *POLG* variants or combinations of heterozygous *POLG* variants classified as pathogenic or likely pathogenic for *POLG*-related mitochondrial disorders (Table 1). Two had homozygous genotypes; the other three had potentially compound heterozygous genotypes for which phase could not be determined.

**Table 1:** Carriers of potentially biallelic pathogenic or likely pathogenic *POLG* variants identified across ancestries.

|  | GP2-ID-1 | GP2-ID-2 | GP2-ID-3 | GP2-ID-4 | GP2-ID-5 |
|---|---|---|---|---|---|
| Genetic Details |  |  |  |  |  |
| *POLG variants* | chr15:89323460:C:G<br>(p.Gly737Arg)<sup>#</sup> | 1) chr15:89329055:A:C<br>(p.Leu304Arg)<sup>#</sup><br>2) chr15:89330184:G:A<br>(p.Thr251Ile)<sup>#</sup><br>3) chr15:89325639:G:A<br>(p.Pro587Leu)<sup>#</sup> | 1) chr15:89323460:C:G<br>(p.Gly737Arg)<sup>#</sup><br>2) chr15:89333327:G:A<br>(p.Ala143Val)<sup>#</sup> | 1) chr15:89325456:G:C<br>(p.Pro648Arg)<sup>#</sup><br>2) chr15:89327201:C:T<br>(p.Ala467Thr)<sup>#</sup> | 1) chr15:89330184:G:A<br>(p.Thr251Ile)<sup>#</sup><br>2) chr15:89325639:G:A<br>(p.Pro587Leu)<sup>#</sup> |
| Zygosity | Hom | Potential compound het | Potential compound het | Potential compound het | Double hom |
| Pathogenicity (ClinVar) | Pathogenic/Likely Pathogenic | 1) Pathogenic/Likely Pathogenic<br>2) + 3) Pathogenic | 1) + 2) Pathogenic/Likely Pathogenic | 1) + 2) Pathogenic/Likely Pathogenic | 1) + 2) Pathogenic |
| Genetic screening method | CES | CES | CES | WGS | CES |
| Other genetic findings | NA | NA | NA | NA | *PRKN: c.1286-3C>G (Hom); pathogenic* |
| Basic Demographics |  |  |  |  |  |
| Genetic ancestry | EUR | EUR | EUR | EUR | AMR |
| Gender | Male | Male | Female | Male | Male |
| Age at sample collection, years | 63 | 45 | 71 | 51 | 42 |
| Clinical Characteristics |  |  |  |  |  |
| Age at onset, years | 55 | 40 | 60 | 46 | 20 |
| Age at most-recent follow-up, years | 63 | 45 | NA | NA | 42 |
| Family history of PD | No | No | Unknown | Yes | Yes |
| Diagnosis | PD | PD | PD | PD | PD |
| Levodopa responsiveness | Unknown | Yes | Unknown | Unknown | Yes |
| Cognition | Unknown | MCI<br>MoCA 23/30 points | Unknown | Unknown | Self-reported MCI<br>MoCA 28/30 points |
| Other comorbidities | Unknown | Kidney disease | Unknown | Unknown | Hypertension |

<sup>#</sup>Previously reported as disease-associated variants under both dominant and recessive inheritance (https://tools.niehs.nih.gov/polg/index.cfm/main/home).
AMR = Latinos and Indigenous People of the Americas, CES = Clinical exome sequencing, EUR = European, Het = Heterozygous, Hom = Homozygous, MCI = Mild cognitive impairment, MoCA = Montreal cognitive assessment,  NA = Not available/applicable.

One EUR PD patient (GP2-ID-1) was homozygous for p.Gly737Arg. Another EUR PD patient (GP2-ID-2) carried the co-occurring *POLG* p.Thr251Ile-p.Pro587Leu variant pair, along with the heterozygous p.Leu304Arg. The third EUR PD case (GP2-ID-3) carried heterozygous p.Gly737Arg and p.Ala143Val variants, whereas the fourth EUR PD case (GP2-ID-4) harboured heterozygous p.Pro648Arg and p.Ala467Thr variants. Lastly, a PD patient of AMR ancestry (GP2-ID-5) was homozygous for both p.Thr251Ile and p.Pro587Leu and also carried a homozygous pathogenic *PRKN* c.1286-3C>G variant. No relationship among these five cases were reported, and no additional pathogenic variants in other genes linked to genetic PD (except GP2-ID-5) were identified.

Clinically, all five were recorded as having typical PD, without documented features suggesting a mitochondrial syndrome, although the available phenotyping was limited and absence of documentation should not be interpreted as absence of symptoms. There was a wide variability in the ages at onset (AAO) of these PD patients, ranging from 20 to 60 years. The youngest case, GP2-ID-5 (onset at 20 years), also had a homozygous pathogenic *PRKN* variant (chr6:161350214:G:C [hg38]; c.1286-3C>G), a variant known to cause early-onset PD, which likely explains the early disease onset in this individual. Among all five carriers, two reported a negative family history of PD, two reported positive history, and one was unknown. Based on the available clinical data, all five patients had a diagnosis of typical PD without mention of relevant atypical features indicating a different diagnosis. Data on medication was only available for two individuals, both of whom showed a good response to dopaminergic treatment. In the context of non-motor features, one individual was found to have mild cognitive impairment (MCI; MoCA score 23/30). Furthermore, two PD cases were found to have comorbidities, including kidney disease and hypertension, respectively. A summary of clinical characteristics is provided in Table 1; overall, the availability of extended clinical data was limited.

### Single heterozygous pathogenic or likely pathogenic *POLG* variant carriers

A total of 34 distinct pathogenic or likely pathogenic variants were reported in heterozygous state across ancestries, most of which were located within the polymerase domain of the POLG protein (Figure 2). A summary of all the variants and their respective frequencies is provided in Table 2; several variants were exclusively identified in PD cases and absent from controls. All variants have been reported before as disease-related for different neurological phenotypes, and the majority of them (n=23/34, 67.6%) have previously been linked to diseases with either dominant or recessive inheritance, according to the National Institute of Environmental Health Sciences (NIEHS) database (https://portal.niehs.nih.gov).

**Figure 1:**
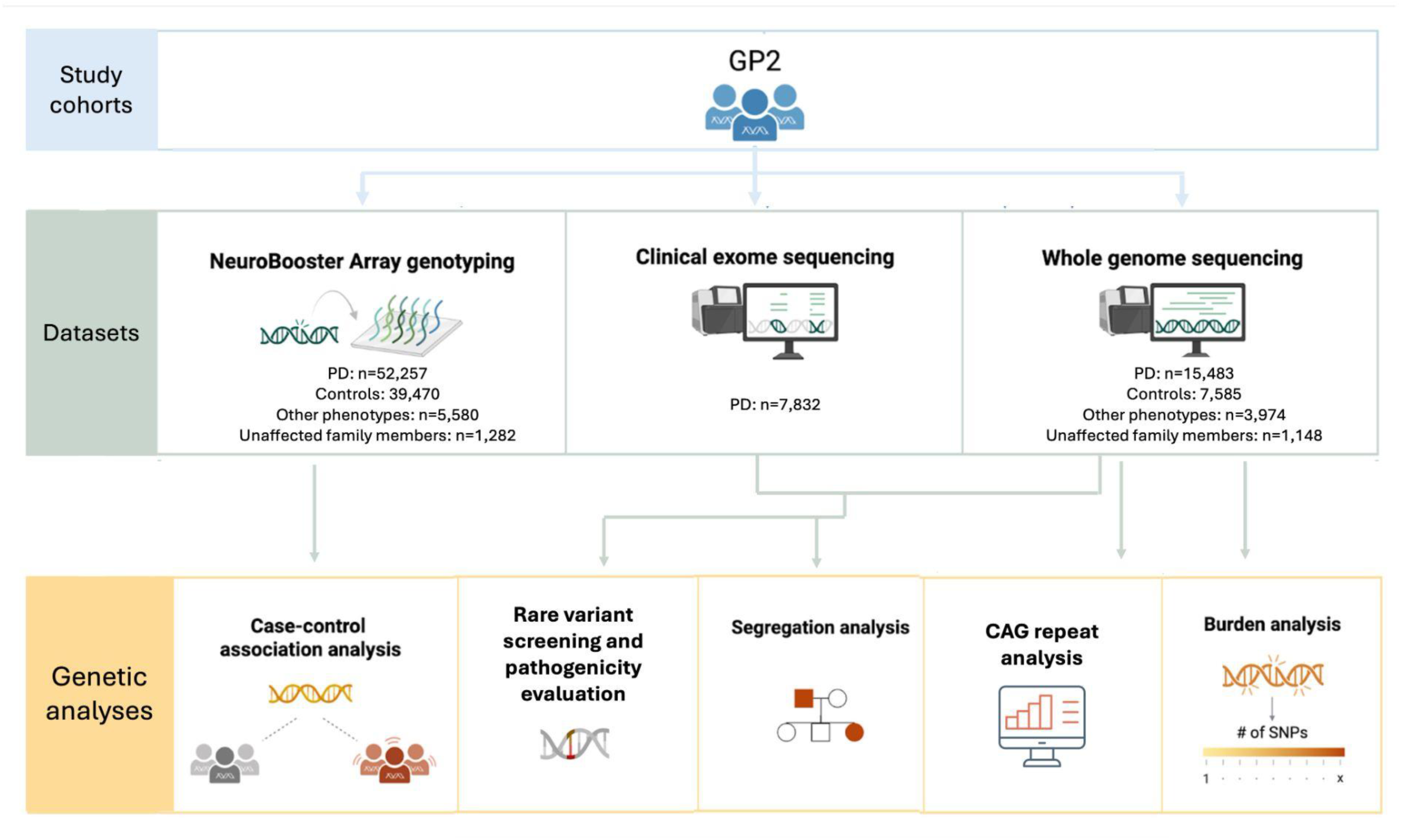
Overview of the study workflow. Data from GP2 release 11 (NBA, CES, and WGS) were leveraged and underwent quality control. Variants in the *POLG* gene were extracted and annotated. Downstream analyses included a case-control association analysis, gene-based burden analysis of rare *POLG* variants, screening for rare *POLG* variants (biallelic and monoallelic), and segregation analysis. CES = Clinical exome sequencing; NBA = NeuroBooster Array genotyping; WGS = Whole genome sequencing.

**Figure 2:**
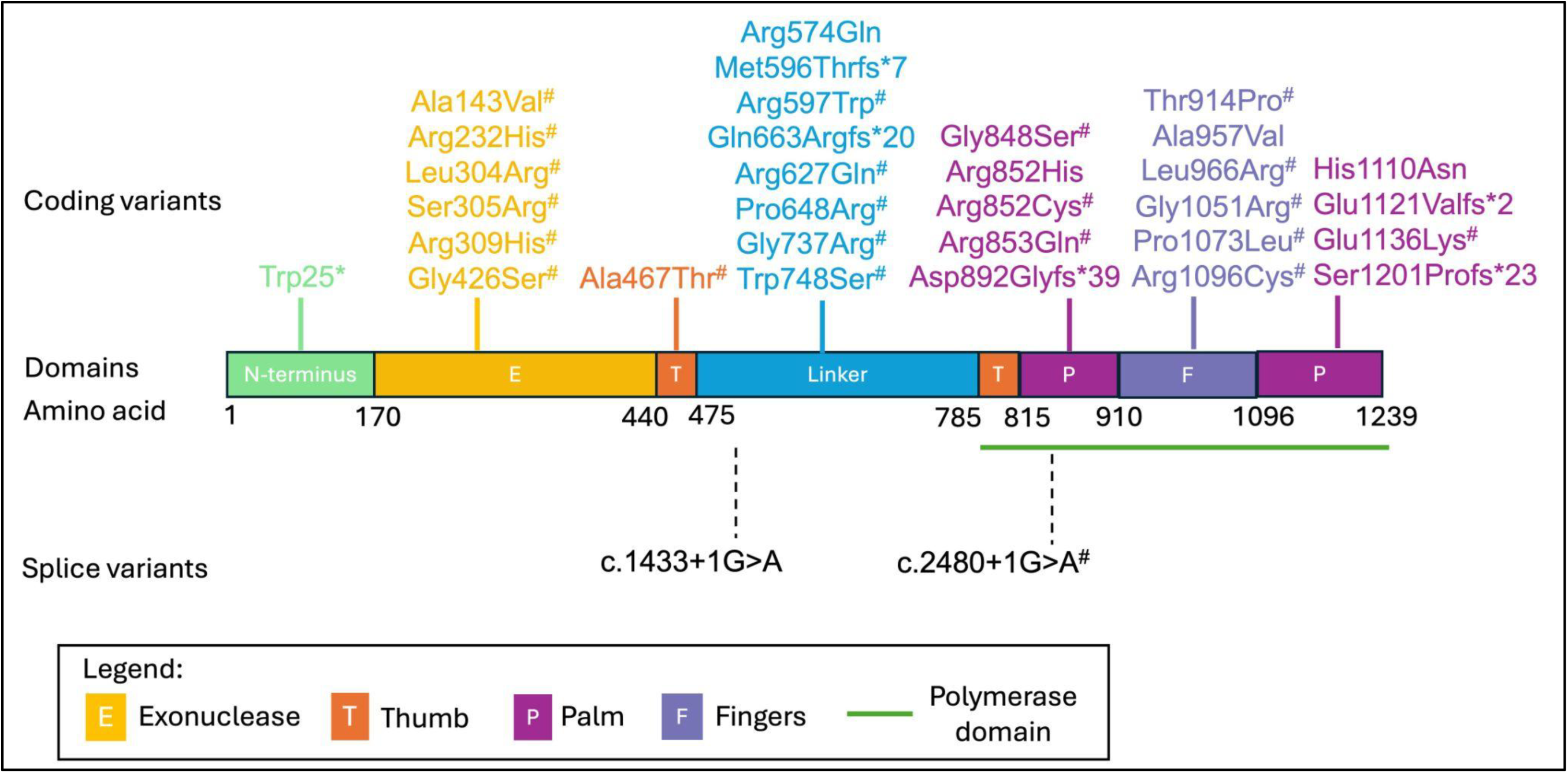
Pathogenic or likely pathogenic *POLG* variants identified across ancestries. Variants predicted to be pathogenic or likely pathogenic were illustrated and mapped onto the domains of POLG protein. The approximate positions of intronic variants, referenced to the protein domains, are indicated by dashed lines. ^#^Previously reported as disease-associated variants under both dominant and recessive inheritance (https://tools.niehs.nih.gov/polg/index.cfm/main/home).

**Table 2:** Carriers of single heterozygous pathogenic or likely pathogenic *POLG* variants across ancestry groups.

| Variant (hg38) | Pathogenicity<br>(ClinVar) | Ancestry | PD<br>(Freq,%) | Other<br>(Freq, %) | Control<br>(Freq, %) | P value<sup>a</sup> | MAF<br>gnomAD<sup>b</sup> |
|---|---|---|---|---|---|---|---|
| chr15:89333680:C:T<br>(p.Trp25*) | P | EUR | 0/8044<br>(0) | 0/3264<br>(0) | 1/2108<br>(0.05) | NA | 8.68E-07 |
| chr15:89333327:G:A<br>(p.Ala143Val)<sup>#</sup> | P/LP | CAS | 1/663<br>(0.15) | 0/165<br>(0) | 0/957<br>(0) | NA | NA |
|  |  | EUR | 4/8044<br>(0.05) | 0/3264<br>(0) | 1/2108<br>(0.05) | 1 | 0.00004338 |
| chr15:89330241:C:T<br>(p.Arg232His)<sup>#</sup> | P | EAS | 0/2697<br>(0) | 0/211<br>(0) | 1/1115<br>(0.09) | NA | 0 |
| chr15:89329055:A:C<br>(p.Leu304Arg)<sup>#</sup> | P/LP | CAH | 1/177<br>(0.56) | 0/50<br>(0) | 0/128<br>(0) | NA | NA |
|  |  | CAS | 1/663<br>(0.15) | 0/165<br>(0) | 0/957<br>(0) | NA | NA |
|  |  | EUR | 2/8044<br>(0.02) | 0/3264<br>(0) | 1/2108<br>(0.05) | 0.5026 | 0.000005085 |
| chr15:89329051:G:C<br>(p.Ser305Arg)<sup>#</sup> | P/LP | EUR | 0/8044<br>(0) | 0/3264<br>(0) | 1/2108<br>(0.05) | NA | 0.000007627 |
| chr15:89329040:C:T<br>(p.Arg309His)<sup>#</sup> | P | EUR | 1/8044<br>(0.01) | 0/3264<br>(0) | 0/2108<br>(0) | NA | 0.00001017 |
| chr15:89327324:C:T<br>(p.Gly426Ser)<sup>#</sup> | P/LP | AJ | 1/741<br>(0.13) | 0/114<br>(0) | 0/669<br>(0) | NA | 0.0001351 |
| chr15:89327201:C:T<br>(p.Ala467Thr)<sup>#</sup> | P | AMR | 1/338<br>(0.30) | 0/20<br>(0) | 0/182<br>(0) | NA | 0.00004998 |
|  |  | EUR | 42/8044<br>(0.52) | 2/3264<br>(0.06) | 4/2108<br>(0.19) | **0.0443** | 0.001330 |
| chr15:89325678:C:T<br>(p.Arg574Gln) | P/LP | CAS | 0/663<br>(0) | 0/165<br>(0) | 1/957<br>(0.10) | NA | NA |
|  |  | EUR | 1/8044<br>(0.01) | 0/3264<br>(0) | 0/2108<br>(0) | NA | 0.00004665 |
|  |  | SAS | 0/447<br>(0) | 0/67<br>(0) | 2/360<br>(0.56) | NA | 0.00002196 |
| chr15:89325612:A:ATCTG<br>(p.Met596Thrfs*7) | P | EUR | 1/8044<br>(0.01) | 0/3264<br>(0) | 0/2108<br>(0) | NA | NA |
| chr15:89325610:G:A<br>(p.Arg597Trp)<sup>#</sup> | P/LP | AFR | 1/1469<br>(0.07) | 0/38<br>(0) | 1/2097<br>(0.05) | 1 | 0.00007996 |
|  |  | EAS | 0/2697<br>(0) | 0/211<br>(0) | 1/1115<br>(0.09) | NA | 0 |
|  |  | EUR | 1/8044<br>(0.01) | 0/3264<br>(0) | 0/2108<br>(0) | NA | 0.000002542 |
| chr15:89324187:CCT:C<br>(p.Gln663Argfs*20) | P/LP | AJ | 0/741<br>(0) | 0/114<br>(0) | 1/669<br>(0.15) | NA | 0 |
|  |  | EUR | 1/8044<br>(0.01) | 0/3264<br>(0) | 0/2108<br>(0) | NA | 0.000001695 |
| chr15:89325519:C:T<br>(p.Arg627Gln)<sup>#</sup> | P/LP | EUR | 0/8044<br>(0) | 1/3264<br>(0.03) | 2/2108<br>(0.09) | NA | 0.00003051 |
| chr15:89325456:G:C<br>(p.Pro648Arg)<sup>#</sup> | P/LP | EUR | 1/8044<br>(0.01) | 0/3264<br>(0) | 0/2108<br>(0) | NA | 0.00001104 |
| chr15:89323460:C:G<br>(p.Gly737Arg)<sup>#</sup> | P/LP | AAC | 0/190<br>(0) | 1/13<br>(7.69) | 0/129<br>(0) | NA | 0.0004263 |
|  |  | AMR | 1/338<br>(0.30) | 0/20<br>(0) | 0/182<br>(0) | NA | 0.0002833 |
|  |  | EUR | 45/8044<br>(0.56) | 13/3264<br>(0.40) | 10/2108<br>(0.47) | 0.7403 | 0.002029 |
| chr15:89323426:C:G<br>(p.Trp748Ser)<sup>#</sup> | P/LP | AMR | 1/338<br>(0.30) | 0/20<br>(0) | 0/182<br>(0) | NA | 0.0001333 |
|  |  | CAS | 0/663<br>(0) | 1/165<br>(0.61) | 0/957<br>(0) | NA | NA |
|  |  | EUR | 12/8044<br>(0.15) | 2/3264<br>(0.06) | 2/2108<br>(0.09) | 0.7479 | 0.0006115 |
|  |  | SAS | 1/447<br>(0.22) | 0/67<br>(0) | 0/360<br>(0) | NA | 0.00005491 |
| chr15:89321792:C:T<br>(p.Gly848Ser)<sup>#</sup> | P | AFR | 0/1469<br>(0) | 0/38<br>(0) | 3/2097<br>(0.14) | NA | 0.0001866 |
|  |  | EUR | 13/8044<br>(0.16) | 6/3264<br>(0.18) | 2/2108<br>(0.09) | 0.7504 | 0.0003941 |
| chr15:89321779:C:T<br>(p.Arg852His) | LP | EAS | 1/2697<br>( 0.04) | 0/211<br>(0) | 0/1115<br>(0) | NA | 0 |
| chr15:89321780:G:A<br>(p.Arg852Cys)<sup>#</sup> | P | EUR | 5/8044<br>(0.06) | 1/3264<br>(0.03) | 2/2108<br>(0.09) | 0.6402 | 0.0001424 |
| chr15:89321776:C:T<br>(p.Arg853Gln)<sup>#</sup> | P/LP | EUR | 1/8044<br>(0.01) | 0/3264<br>(0) | 0/2108<br>(0) | NA | 0.000002542 |
| chr15:89321184:T:TC<br>(p.Asp892Glyfs*39) | P | EUR | 1/8044<br>(0.01) | 0/3264<br>(0) | 0/2108<br>(0) | NA | 0.00000339 |
| chr15:89321007:T:G<br>(p.Thr914Pro)<sup>#</sup> | P/LP | EUR | 3/8044<br>(0.04) | 0/3264<br>(0) | 0/2108<br>(0) | NA | 0.00025 |
| chr15:89320877:G:A<br>(p.Ala957Val) | P/LP | EUR | 1/8044<br>(0.01) | 0/3264<br>(0) | 0/2108<br>(0) | NA | 8.47E-07 |
| chr15:89320850:A:C<br>(p.Leu966Arg)<sup>#</sup> | P/LP | EUR | 1/8044<br>(0.01) | 1/3264<br>(0.03) | 0/2108<br>(0) | NA | 0.00005678 |
| chr15:89319053:C:G<br>(p.Gly1051Arg)<sup>#</sup> | P/LP | EUR | 2/8044<br>(0.02) | 0/3264<br>(0) | 1/2108<br>( 0.05) | 0.5026 | 0.00004153 |
|  |  | MDE | 2/694<br>(0.29) | 0/26<br>(0) | 0/984<br>(0) | NA | 0.0004949 |
| chr15:89319053:C:T<br>(p.Gly1051Arg)<sup>#</sup> | P/LP | EAS | 1/2697<br>(0.04) | 0/211<br>(0) | 0/1115<br>(0) | NA | 0 |
| chr15:89318986:G:A<br>(p.Pro1073Leu)<sup>#</sup> | P | AFR | 1/1469<br>(0.07) | 0/38<br>(0) | 0/2097<br>(0) | NA | 0 |
|  |  | EAS | 2/2697<br>(0.07) | 0/211<br>(0) | 0/1115<br>(0) | NA | 0.00002228 |
| chr15:89318737:G:A<br>(p.Arg1096Cys)<sup>#</sup> | LP | CAS | 0/663<br>(0) | 0/165<br>(0) | 1/957<br>(0.10) | NA | NA |
|  |  | MDE | 1/694<br>(0.14) | 0/26<br>(0) | 0/984<br>(0) | NA | 0 |
| chr15:89318695:G:T<br>(p.His1110Asn) | VUS/LP | EUR | 1/8044<br>(0.01) | 0/3264<br>(0) | 0/2108<br>(0) | NA | NA |
| chr15:89318661:T:TCAAA<br>(p.Glu1121Valfs*2) | P/LP | EUR | 1/8044<br>(0.01) | 0/3264<br>(0) | 0/2108<br>(0) | NA | 8.474e-7 |
| chr15:89318617:C:T<br>(p.Glu1136Lys)<sup>#</sup> | P/LP | EUR | 1/8044<br>(0.01) | 0/3264<br>(0) | 0/2108<br>(0) | NA | 0.000002542 |
| chr15:89317417:GA:G<br>(p.Ser1201Profs*23) | P/LP | EUR | 1/8044<br>(0.01) | 0/3264<br>(0) | 0/2108<br>(0) | NA | 0.00000339 |
| chr15:89327166:C:T<br>(c.1433+1G>A) | P | AFR | 1/1469<br>(0.07) | 0/38<br>(0) | 1/2097<br>(0.05) | 1 | 0.0001468 |
| chr15:89321961:C:T<br>(c.2480+1G>A)<sup>#</sup> | P | EAS | 1/2697<br>(0.04) | 0/211<br>(0) | 0/1115<br>(0) | NA | 0 |

<sup>a</sup> Fisher’s exact test was used to compare the frequency of the *POLG* variants among PD cases and controls. Carrier counts and frequencies reflect combined data from WGS and CES cohorts.
<sup>b</sup> For each variant, we reported the ancestry-specific minor allele frequency (MAF) based on data from gnomAD v4.1.0. Frequencies of ancestry groups in our study without corresponding groups available on gnomAD are labeled as NA.
<sup>#</sup>Previously reported as disease-associated variants under both dominant and recessive inheritance (https://tools.niehs.nih.gov/polg/index.cfm/main/home).
AAC = African Admixed, AFR = African, AJ = Ashkenazi Jewish, AMR = Latinos and Indigenous People of the Americas, CAH = Complex Admixture History, CAS = Central Asian, CES = clinical exome sequencing, EAS = East Asian, EUR = European, Freq = Frequency, FIN = Finnish, MAF = minor allele frequency, LP = Likely pathogenic, MDE = Middle Eastern, NA = Not available/applicable, P = Pathogenic, SAS = South Asian, VUS = Variant of uncertain significance, WGS = Whole genome sequencing.

Of the 34 variants, two were found in members of two European families. The p.Gly737Arg variant was identified in two siblings, both affected with PD (AAO 64 and 69 years, respectively) supporting potential familial cosegregation. In the second family, p.Ala467Thr was found in a PD proband with a very early disease onset (AAO 18 years) and his unaffected mother (aged 67 years). Notably, the early-onset PD patient also carried a *PRKN* deletion involving exons 3-4 and the p.Cys253Tyr missense variant, whereas his unaffected mother carried only the *PRKN* missense variant. Overall, heterozygous pathogenic or likely pathogenic *POLG* variants were present in <1% of PD patients (n=161/23,315; 0.69%), patients with other neurological phenotypes (n=28/3,974; 0.70%), and controls (n=39/7,585; 0.51%) across all ancestries. No significant enrichment was observed in PD patients as compared to controls (OR=1.35, 95% CI=0.94-1.96, p=0.0996). At the individual variant level, p.Ala467Thr showed nominal enrichment in PD patients of European ancestry (p=0.0443; Table 2).

### Carrier frequency of two co-occurring *POLG* variants

The *POLG* variants p.Thr251Ile and p.Pro587Leu variants have been reported together on a cis haplotype and are classified as pathogenic in combination according to ClinVar (Supplementary Figure 1). We examined their co-occurrence among PD patients, individuals with other phenotypes, and controls in the NBA and WGS datasets. Carrier frequencies were below 1% in the reported ancestry groups, with no statistically significant case-control differences (Table 3).

**Table 3:** Carrier frequency of the co-inherited *POLG* p.Thr251Ile and p.Pro587Leu variants across ancestries.

| Ancestry | NBA |  |  |  | WGS |  |  |  |
|---|---|---|---|---|---|---|---|---|
|  | PD (Freq,%) | Other (Freq,%) | Control (Freq,%) | P value<sup>a</sup> | PD (Freq,%) | Other (Freq,%) | Control (Freq,%) | P value<sup>a</sup> |
| AAC | 1/491<br>(0.20) | 0/18<br>(0.00) | 1/846<br>(0.12) | 1.000 | 0/190<br>(0.00) | 0/13<br>(0.00) | 0/129<br>(0.00) | NA |
| AFR | 0/2770<br>(0.00) | 0/44<br>(0.00) | 1/4412<br>(0.02) | NA | 0/1469<br>(0.00) | 0/38<br>(0.00) | 0/2097<br>(0.00) | NA |
| AJ | 3/2225<br>(0.13) | 0/29<br>(0.00) | 0/801<br>(0.00) | NA | 2/741<br>(0.27) | 0/114<br>(0.00) | 4/669<br>(0.60) | 0.4314 |
| AMR | 7/2222<br>(0.32) | 0/28<br>(0.00) | 0/1508<br>(0.00) | NA | 0/338<br>(0.00) | 0/20<br>(0.00) | 0/182<br>(0.00) | NA |
| CAH | NA | NA | NA | NA | 1/177<br>(0.56) | 0/50<br>(0.00) | 0/128<br>(0.00) | NA |
| CAS | 2/1194<br>(0.17) | 0/175<br>(0.00) | 1/1373<br>(0.07) | 0.6006 | 1/663<br>(0.15) | 0/165<br>(0.00) | 0/957<br>(0.00) | NA |
| EAS | 0/4609<br>(0.00) | 0/396<br>(0.00) | 0/2769<br>(0.00) | NA | 0/2697<br>(0.00) | 0/211<br>(0.00) | 0/1115<br>(0.00) | NA |
| EUR | 129/35955<br>(0.36) | 15/4625<br>(0.32) | 96/25598<br>(0.38) | 0.7354 | 67/8044<br>(0.83) | 19/3264<br>(0.58) | 14/2108<br>(0.66) | 0.4940 |
| FIN | 0/113<br>(0.00) | 0/11<br>(0.00) | 0/19<br>(0.00) | NA | 0/23<br>(0.00) | 0/6<br>(0.00) | 0/4<br>(0.00) | NA |
| MDE | 2/1100<br>(0.18) | 1/35<br>(2.86) | 2/1304<br>(0.15) | 1.000 | 2/694<br>(0.29) | 0/26<br>(0.00) | 4/984<br>(0.41) | 1.000 |
| SAS | 0/739<br>(0.00) | 0/103<br>(0.00) | 0/398<br>(0.00) | NA | 0/447<br>(0.00) | 0/67<br>(0.00) | 0/360<br>(0.00) | NA |

<sup>a</sup> Fisher’s exact test was used to compare the frequency of the two *POLG* variants among PD cases and controls. Comparison was not available when no event was observed in either group.
AAC = African Admixed, AFR = African, AJ = Ashkenazi Jewish, AMR = Latinos and Indigenous People of the Americas, CAH = Complex Admixture History, CAS = Central Asian, EAS = East Asian, EUR = European, Freq = Frequency, FIN = Finnish, NBA = NeuroBooster Array, MDE = Middle Eastern, NA = Not available/applicable, SAS = South Asian, WGS = Whole genome sequencing.

### *POLG* CAG repeat length distribution

The 10/10 CAG repeat genotype was the most frequently observed genotype across all ancestry groups, although its frequency varied substantially between populations (Table 4). Among PD patients, the frequency of the 10/10 genotype ranged from 42.1% in African admixed (AAC) individuals and 44.4% in Africans (AFR) to 94.6% in East Asians, with similar patterns observed among controls. The 10/x and x/x genotypes were less frequent across most ancestry groups, although the x/x genotypes were more prevalent among AFR (10.9%) and AAC (7.9%) PD patients, as well as among controls (11.6% and 4.6%, respectively). Overall, genotype distributions were broadly comparable between PD patients and controls within ancestry groups.

**Table 4:** Distribution of *POLG* CAG repeat genotypes and their association with Parkinson’s disease risk across ancestry groups.

| **Ancestry** | **PD, n (%)** |  |  | **Other, n (%)** |  |  | **Control, n (%)** |  |  | **OR (95% CI); p<sup>a</sup>** |  |  |
|---|---|---|---|---|---|---|---|---|---|---|---|---|
| **Genotypes** | **10/10** | **10/x** | **x/x** | **10/10** | **10/x** | **x/x** | **10/10** | **10/x** | **x/x** | **Non-10/10 vs. 10/10** | **Non-11/11 vs. 11/11** | **Non-10/11 vs. 10/11** |
| AAC | 80 (42.1) | 95 (50.0) | 15 (7.9) | 2 (40.0) | 2 (40.0) | 1 (20.0) | 75 (49.7) | 69 (45.7) | 7 (4.6) | **1.84 (1.06–3.18); 0.0291** | 1.76 (0.35–8.81); 0.4894 | 0.71 (0.39–1.29); 0.2570 |
| AFR | 581 (44.4) | 585 (44.7) | 142 (10.9) | 14 (43.8) | 13 (40.6) | 5 (15.6) | 890 (44.7) | 872 (43.8) | 231 (11.6) | 1.02 (0.88–1.19); 0.7524 | 0.92 (0.66–1.28); 0.6254 | 1.01 (0.85–1.19); 0.9250 |
| AJ | 730 (71.5) | 268 (26.2) | 23 (2.3) | 36 (80.0) | 9 (20.0) | 0 (0.0) | 193 (75.7) | 58 (22.7) | 4 (1.6) | 1.23 (0.83–1.81); 0.3074 | 0.82 (0.18–3.71); 0.7984 | 0.84 (0.54–1.31); 0.4524 |
| AMR | 584 (75.7) | 166 (21.5) | 21 (2.7) | 3 (75.0) | 0 (0.0) | 1 (25.0) | 247 (80.7) | 54 (17.6) | 5 (1.6) | 1.18 (0.80–1.76); 0.4019 | 0.67 (0.07–6.28); 0.7286 | 0.83 (0.47–1.45); 0.5035 |
| CAH<sup>#</sup> | 126 (67.7) | 54 (29.0) | 6 (3.2) | 17 (68.0) | 8 (32.0) | 0 (0.0) | 84 (68.3) | 34 (27.6) | 5 (4.1) | 1.07 (0.61–1.88, 0.8236) | NA | 0.94 (0.46–1.95, 0.8762) |
| CAS<sup>#</sup> | 552 (86.5) | 81 (12.7) | 5 (0.8) | 140 (89.7) | 15 (9.6) | 1 (0.6) | 825 (84.9) | 141 (14.5) | 6 (0.6) | 1.00 (0.73–1.35, 0.9771) | 1.98 (0.22–17.41, 0.5394) | 0.96 (0.67–1.37, 0.8285) |
| EAS | 1462 (94.6) | 83 (5.4) | 1 (0.1) | 436 (97.3) | 12 (2.7) | 0 (0.0) | 951 (92.1) | 80 (7.7) | 2 (0.2) | 0.70 (0.49–1.01); 0.0570 | NA | 1.27 (0.81–1.99); 0.2996 |
| EUR<sup>#</sup> | 5326 (70.3) | 2061 (27.2) | 193 (2.5) | 1671 (70.6) | 624 (26.4) | 71 (3.0) | 893 (72.5) | 301 (24.4) | 38 (3.1) | 1.09 (0.95–1.25, 0.2330) | 1.01 (0.59–1.73, 0.9731) | 0.96 (0.81–1.13, 0.5978) |
| MDE<sup>#</sup> | 589 (73.3) | 201 (25.0) | 13 (1.6) | 16 (69.6) | 6 (26.1) | 1 (4.3) | 682 (70.7) | 253 (26.2) | 30 (3.1) | 0.99 (0.77–1.26, 0.9093) | 1.83 (0.75–4.47, 0.1817) | 0.96 (0.73–1.27, 0.7783) |
| SAS | 219 (75.8) | 67 (23.2) | 3 (1.0) | 41 (69.5) | 15 (25.4) | 3 (5.1) | 245 (71.4) | 92 (26.8) | 6 (1.7) | 0.72 (0.48–1.09); 0.1210 | 2.47 (0.43–14.04); 0.3077 | 1.31 (0.83–2.07); 0.2394 |

<sup>#</sup>Logistic regression models were adjusted for age, sex, and the first five genetic principal components (PCs) for all ancestries except in the CAH, CAS, EUR, and MDE, which were adjusted only for sex and the first five PCs due to substantial missingness in age.

Findings from the genotype association analyses between *POLG* CAG repeat genotypes and PD risk across ancestries are summarized in Table 4. In the non-10/10 versus 10/10 comparison, a nominally significant association was observed in the AAC cohort (OR=1.84, 95% CI=1.06-3.18, p=0.0291), but failed to survive the Bonferroni correction. For the non-11/11 versus 11/11 and non-10/11 versus 10/11 comparisons, no nominal significance was observed. A detailed breakdown of the genotypes identified in each ancestry was summarized in Supplementary Table 2 and forest plots illustrating the distribution of repeat genotypes were shown in Figure 3.

**Figure 3:**
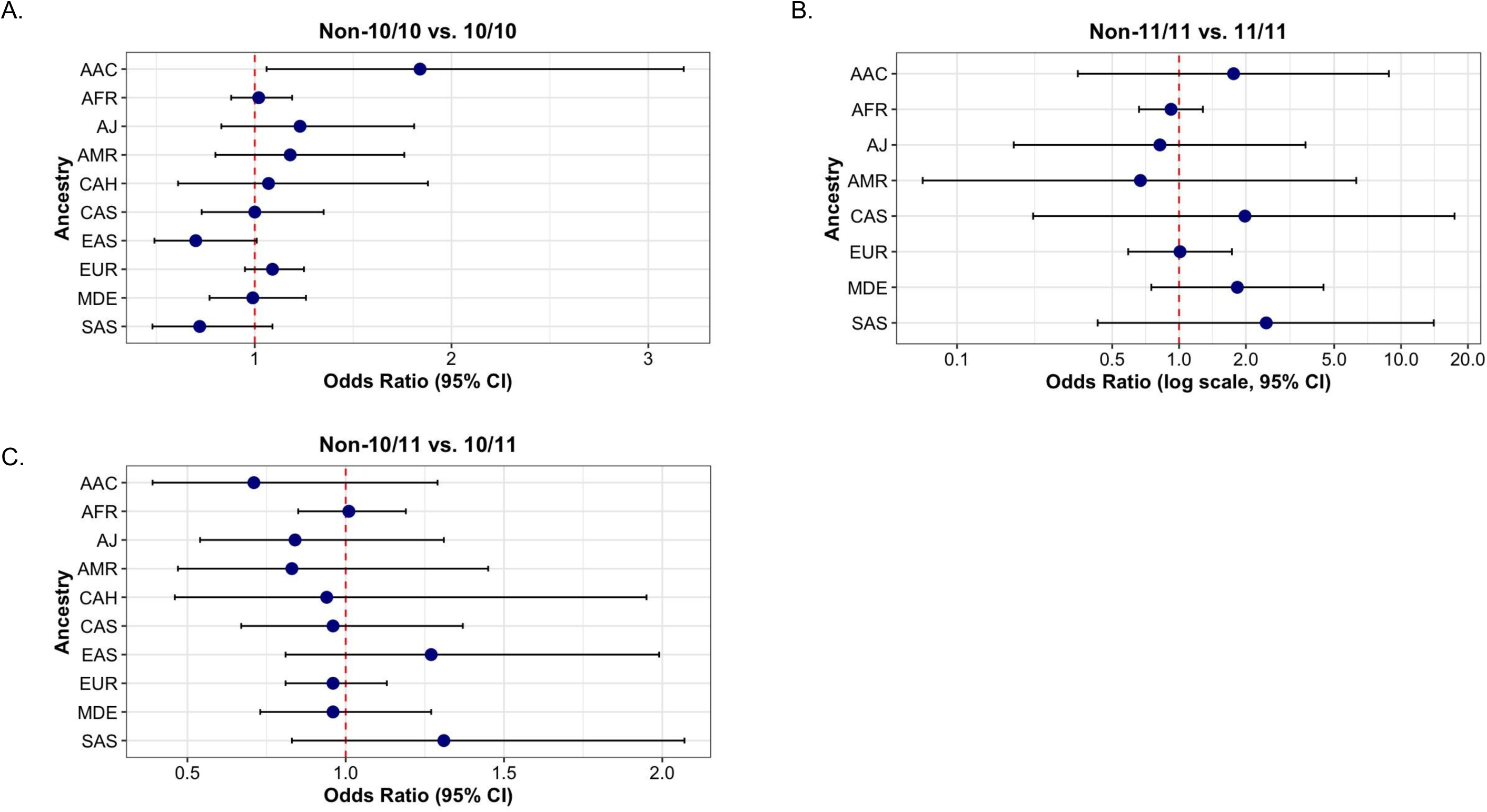
Forest plots of *POLG* repeat genotypes across ancestries. AAC = African Admixed, AFR = African, AJ = Ashkenazi Jewish, AMR = Latinos and Indigenous People of the Americas, CAH = Complex Admixture History, CAS = Central Asian, CI = Confidence interval, EAS = East Asian, EUR = European, FIN = Finnish, NBA = NeuroBooster Array, MDE = Middle Eastern, SAS = South Asian.

### Gene-based rare variant analysis

The results of the burden analysis across the investigated variant masks and ancestries are summarized in Supplementary Table 3. However, none of the masks showed significant association with PD risk.

### Case-control analysis of common *POLG* variants

Of the 16 common (MAF ≥1%) exonic *POLG* variants retained after quality control, only one showed a nominal association with PD risk: the synonymous variant p.Leu752= (chr15-89323415 G>A; rs41564016) in individuals of EUR ancestry (p=0.0418). However, this association did not survive Bonferroni correction. A summary of all exonic *POLG* variants is provided in Supplementary Table 4, and regional association plots for each ancestry group are shown in Supplementary Figure 2.

## Discussion

In this study, we characterized the frequency and spectrum of *POLG* variants in individuals with PD, other neurological phenotypes, and unaffected controls across diverse genetic ancestries. To our knowledge, this represents the largest multi-ancestry analysis to date examining the relationship between *POLG* genetic variation and PD. By integrating rare pathogenic and likely pathogenic variants, common genetic variation, and *POLG* CAG repeat genotypes, we found no robust evidence supporting an association between *POLG* variation and PD risk after correction for multiple testing. Nevertheless, we identified a small number of individuals with PD carrying homozygous or potentially compound heterozygous *POLG* variants, highlighting the possibility that *POLG* may contribute to PD in a small number of individuals.

Five PD patients carried homozygous or multiple heterozygous *POLG* variants previously associated with mitochondrial disorders. Among these, GP2-ID-1 was homozygous for p.Gly737Arg, whereas GP2-ID-3 carried p.Gly737Arg and p.Ala143Val, although the phase of these variants could not be established. The p.Gly737Arg variant has previously been implicated in mitochondrial and peripheral nerve disorders, with clinical manifestations including ataxia, seizures, neuropathy, and parkinsonism.^3^ It has been reported in compound heterozygosity with other *POLG* variants (e.g., p.Arg309His, p.Ser64Leu), and associated with axonal Charcot-Marie-Tooth-like neuropathy, occasional dystonia, and parkinsonism features.^36–38^ Notably, two sisters with early-onset PD and sensory neuropathy were previously reported to carry the p.Gly737Arg and p.Arg853Trp variants on different haplotypes (*trans*).^8^ Although these observations support the biological plausibility of *POLG*-related mitochondrial dysfunction contributing to parkinsonism, the absence of detailed clinical phenotyping and segregation data in our cohort prevents conclusions regarding the pathogenic contribution of these variants to the observed PD phenotypes.

The co-occurring *POLG* p.Thr251Ile and p.Pro587Leu variants were identified together with p.Leu304Arg in GP2-ID-2 and in the homozygous state in GP2-ID-5. These variants have been extensively described as a pathogenic cis haplotype in mitochondrial disorders,^39–42^ occurring either in homozygosity,^42^ or in combination with additional *POLG* variants.^43,44^ Functional studies have demonstrated that, although each variant individually produces a relatively modest reduction in polymerase γ activity, their co-occurrence on the same allele acts synergistically, reducing DNA-binding affinity and polymerase activity to approximately 5-10% of wild-type levels.^33^ Clinically, this haplotype has been associated with PEO and multisystemic mitochondrial manifestations, including ataxia, myopathy, epilepsy, neuropathy, and hepatic dysfunction.^33,41^

Clinically, all five individuals carrying homozygous or potentially compound heterozygous *POLG* variants had been diagnosed with typical PD, without obvious atypical features and with a good response to dopaminergic treatment in two individuals with available medication data. The ages at onset varied substantially, ranging from 20 to 60 years. Notably, one individual also carried a homozygous pathogenic *PRKN* variant, which is more likely to represent the disease-causing alteration in this case and is consistent with the very early onset of 20 years observed in this individual. While additional clinical data was very limited, particularly on non-motor features, one individual showed mild cognitive impairment. Interestingly, cognitive dysfunction has been previously reported in *POLG*-related disorders, including *POLG*-associated parkinsonism,^41,44^ although its frequency and severity appear to vary depending on the disease phenotype, age at onset, and underlying variant combination. Unfortunately, additional clinical information on variant carriers, including potential mitochondrial and systemic features, were unavailable, precluding us from investigating potential genotype-phenotype correlations in more detail. Based on the limited clinical data, the small number of affected individuals, and the uncertainty regarding true biallelic status due to the constraints of the available sequencing data, we cannot conclusively determine whether the identified *POLG* variants are causal, contributory, or merely coincidental in relation to the PD phenotype.

Among individuals carrying single heterozygous pathogenic or likely pathogenic *POLG* variants, we observed an enrichment of p.Ala467Thr in PD patients of European ancestry (p=0.043). This variant has been reported to occur in ∼0.6% of the Belgian population,^45^ and is one of the most frequently detected *POLG* variants associated with Alpers syndrome, autosomal recessive PEO and SANDO.^46^ The variant is located within the linker region and has been found to reduce the catalytic efficiency of POLG via *in vitro* study.^47^ Nevertheless the observed association with PD was nominal and should be interpreted cautiously. In addition, we identified two families with multiple *POLG* variant carriers. The Gly737Arg was present in two siblings diagnosed with PD at ages 64 and 69 years, respectively, without additional identified pathogenic variants in established PD genes. Although this observation raises the possibility of a contribution to disease susceptibility, the absence of extended segregation data, unaffected family members, and functional validation prevents conclusions regarding its pathogenic role in the heterozygous state. In another family, p.Ala467Thr was identified in an individual with very early-onset PD (18 years) and his unaffected mother, aged 67 years. Importantly, the affected individual also carried a *PRKN* deletion involving exons 3-4 and the p.Cys253Tyr variant, potentially driving an alternative genetic explanation for his phenotype. However, confirmation of the phase is necessary before assigning causality. Taken together, these familial observations illustrate the difficulties of interpreting heterozygous *POLG* variants in PD, particularly in the presence of additional genetic findings and without comprehensive segregation information.

Overall, the role of heterozygous *POLG* variants in PD remains uncertain. While *POLG* is classically more commonly linked to recessive mitochondrial diseases, single pathogenic variants may act as reduced-penetrance risk alleles or require an additional genetic, environmental, or age-related “second hit”. This has important implications for clinical interpretation and particularly genetic counseling, as the presence of a single pathogenic *POLG* variant alone does not necessarily establish causality for PD. Future studies should therefore incorporate the systematic assessment of mitochondrial features to better estimate pathogenicity and distinguish incidental carrier status from *POLG*-related disease presenting with parkinsonism. Detailed phenotyping will be particularly important for counseling carriers, where current evidence often does not allow a clear distinction between causal, contributory, and incidental findings.

We also investigated the co-inherited p.Thr251Ile-p.Pro587Leu haplotype across genetic ancestries. Carrier frequencies were below 1% among both PD cases and controls, with no statistically significant differences between groups. The frequency observed in controls was broadly consistent with previous observations in an Italian population,^48,49^ where this haplotype was reported in approximately 1% of individuals. To our knowledge, our study is the first to examine the frequency of these two co-inherited variants across ancestries and at a large scale. Despite their predicted pathogenicity, the absence of significant enrichment among PD cases suggests that this haplotype is unlikely to represent a major genetic factor relevant to PD.

Variation in the *POLG* exon 2 CAG repeat tract has previously been investigated as a potential genetic risk factor for PD, with inconsistent findings across populations. In our multi-ancestry analysis, the 10/10 genotype was the most frequent across all ancestry groups, although its prevalence varied substantially. Non-10 repeat genotypes were more common among individuals of African and African admixed ancestries^50^ than among several other ancestral groups,^15,22,51,52^ consistent with previously reported population differences in *POLG* CAG repeat variation. Although nominal association was observed for specific genotype comparison in the African admixed ancestry group, it did not survive multiple correction, and the overall directions of association across ancestries were inconsistent. These results do not support a robust association between the investigated *POLG* CAG repeat genotypes and PD susceptibility. They also highlight the importance of accounting for ancestry-specific repeat distributions when evaluating associations in genetically diverse populations.

Consistent with these findings, gene-based rare variant analyses using WGS data did not identify significant associations between *POLG* variation and PD risk after correction for multiple testing. Although a nominal association was observed in the East Asian ancestry group, this finding should be interpreted cautiously given the number of comparisons performed and the limited statistical power to detect rare variant effects in some ancestry groups. Similarly, our single-variant analyses of common *POLG* variants identified only a nominal association for the synonymous variant p.Leu752= in individuals of European ancestry, which did not survive multiple testing corrections. These findings are broadly consistent with previous studies reporting no robust association between common *POLG* variation and idiopathic PD.^15,16,19^

A major strength of this study is the large, genetically diverse sample, which allowed us to investigate multiple classes of *POLG* variation using complementary approaches. The inclusion of individuals from ancestrally historically underrepresented in genetic studies of PD also enabled us to characterize population-specific differences in variant frequencies and CAG repeat distributions. However, several limitations should be acknowledged. First, despite the large overall sample size, the number of carriers of individual pathogenic or likely pathogenic variants was small, and some ancestry groups had limited numbers of cases and controls, reducing statistical power, particularly for rare variant analysis. Second, the phase of multiple heterozygous variants could not be established, limiting the interpretation of potentially compound heterozygous genotypes. Third, the available clinical information was insufficient to systematically assess mitochondrial manifestations or investigate genotype-phenotype relationships. Fourth, pathogenicity classifications are primarily derived from associations with established mitochondrial disorders rather than PD, and therefore may not directly reflect the relevance of individual variants to PD. Finally, individuals presenting with classical *POLG*-related mitochondrial syndromes (e.g., myopathy, PEO, or SANDO) accompanied by parkinsonism may have been underrepresented because GP2 recruitment primarily focuses on individuals diagnosed with PD. Consequently, our findings may not fully capture the contribution of *POLG* variation to the broader spectrum of mitochondrial parkinsonian disorders.

In conclusion, our large-scale multi-ancestry analysis provides no robust evidence supporting an association between *POLG* genetic variation and PD susceptibility at the population level. Although pathogenic or likely pathogenic *POLG* variants were identified in a small proportion of individuals with PD, their frequencies were broadly comparable between cases and controls, and neither rare variant burden nor common variant and CAG repeat analyses revealed significant associations after correction for multiple testing. The identification of patients carrying homozygous or potentially compound heterozygous *POLG* variants highlights the possibility that *POLG* may contribute to parkinsonism in a small number of individuals. However, limited clinical phenotyping, unresolved variant phase, and the presence of alternative genetic explanations in some cases prevent the establishment of a causal relationship. Future studies combining detailed clinical phenotyping, family-based segregation analyses, improved variant characterization, and functional assessment of mitochondrial dysfunction will be essential to clarify the role of *POLG* in parkinsonian phenotypes.

## Acknowledgement

This project was supported by the Global Parkinson’s Genetics Program (GP2; https://gp2.org). GP2 is funded by the Aligning Science Across Parkinson’s (ASAP) initiative and implemented by The Michael J. Fox Foundation for Parkinson’s Research (MJFF). For a complete list of GP2 members see doi.org/10.5281/zenodo.7904831.

This research was supported in part by the Intramural Research Program of the National Institutes of Health (NIH). The contributions of the NIH author(s) are considered Works of the United States Government. The findings and conclusions presented in this paper are those of the author(s) and do not necessarily reflect the views of the NIH or the U.S. Department of Health and Human Services.

## Data and Code Availability

Data used in the preparation of this article were obtained from the Global Parkinson’s Genetics Program (GP2; https://gp2.org). Specifically, we used Tier 2 data from GP2 release 11 (10.5281/zenodo.17753486). GP2 data can be accessed through AMP PD (https://amp-pd.org). All code generated for this article, and the identifiers for all software programs and packages used, are available on GitHub (https://github.com/GP2code/POLG_multiancestry) and were given a persistent identifier via Zenodo (10.5281/zenodo.19861264).

## Author contributions

CMS, LML and MTP conceptualized and designed the study. YWT, IE, DY, MJ, ZHF, LML and MTP performed the data analysis, and contributed to data interpretation together with AHT and CMS. HC, AAD, and RNA contributed samples and expanded clinical data to this study. YWT, IE, LML and MTP drafted the manuscript. All authors critically reviewed and approved the final version of the manuscript.

## Competing interest

YWT, LS, MJ and MTP declare no financial conflict of interest. IE was supported by the H2020-MSCA-COFUND-2019 No. 945425 “DevelopMed”. DY is supported by a PhD scholarship from Muscular Dystrophy NSW. PJK received grants from the National Science and Technology Council (NSTC), Taiwan, unrelated to this manuscript. AAD receives consulting fees from the Parkinson’s Foundation. Unrelated to this manuscript, she receives consulting fees from McGill University. RNA received research funding from the Michael J. Fox Foundation, the Silverstein Foundation, the Parkinson’s Foundation and the Aufzien Family Center for the Prevention and Treatment of Parkinson’s Disease. He received consultation fees from Bexxion, Biogen, Biohaven, Capsida, Gain Therapeutics, Genzyme/Sanofi, Janssen, SK Biopharmaceuticals, Takeda, Teva and Vanqua Bio. ZHF is supported by the Aligning Science Across Parkinson’s (ASAP) Global Parkinson’s Genetics Program (GP2) and received GP2 salary support from The Michael J. Fox Foundation for Parkinson’s Research. AHT receives support from the Michael J Fox Foundation and the Global Parkinson Genetic Program (GP2). Unrelated to this manuscript, she received speaker honoraria from the International Parkinson and Movement Disorders Society, Eisai and Orion Pharma and reports consultancies from Elsevier as Section Editor for Parkinsonism and Related Disorders. CMS receives support from the Michael J Fox Foundation, Shake it up Australia Foundation, the Global Parkinson Genetic Program (GP2), National Health and Medical Research Council of Australia and the Medical Research Future Fund of Australia. Unrelated to this manuscript, she received speaker honoraria from the International Parkinson and Movement Disorders Society, and reports consultancies from Abbvie. LML received faculty honoraria from the International Parkinson and Movement Disorders Society, unrelated to this manuscript.

## Funding

This project was supported by the Global Parkinson’s Genetics Program (GP2; https://gp2.org). GP2 is funded by the Aligning Science Across Parkinson’s (ASAP) initiative and implemented by The Michael J. Fox Foundation for Parkinson’s Research (MJFF). For a complete list of GP2 members see doi.org/10.5281/zenodo.7904831. This research was supported in part by the Intramural Research Program of the National Institutes of Health (NIH). The contributions of the NIH author(s) are considered Works of the United States Government. The findings and conclusions presented in this paper are those of the author(s) and do not necessarily reflect the views of the NIH or the U.S. Department of Health and Human Services.

## Supplementary Material

### Supplementary Figures

**Supplementary Figure 1:**
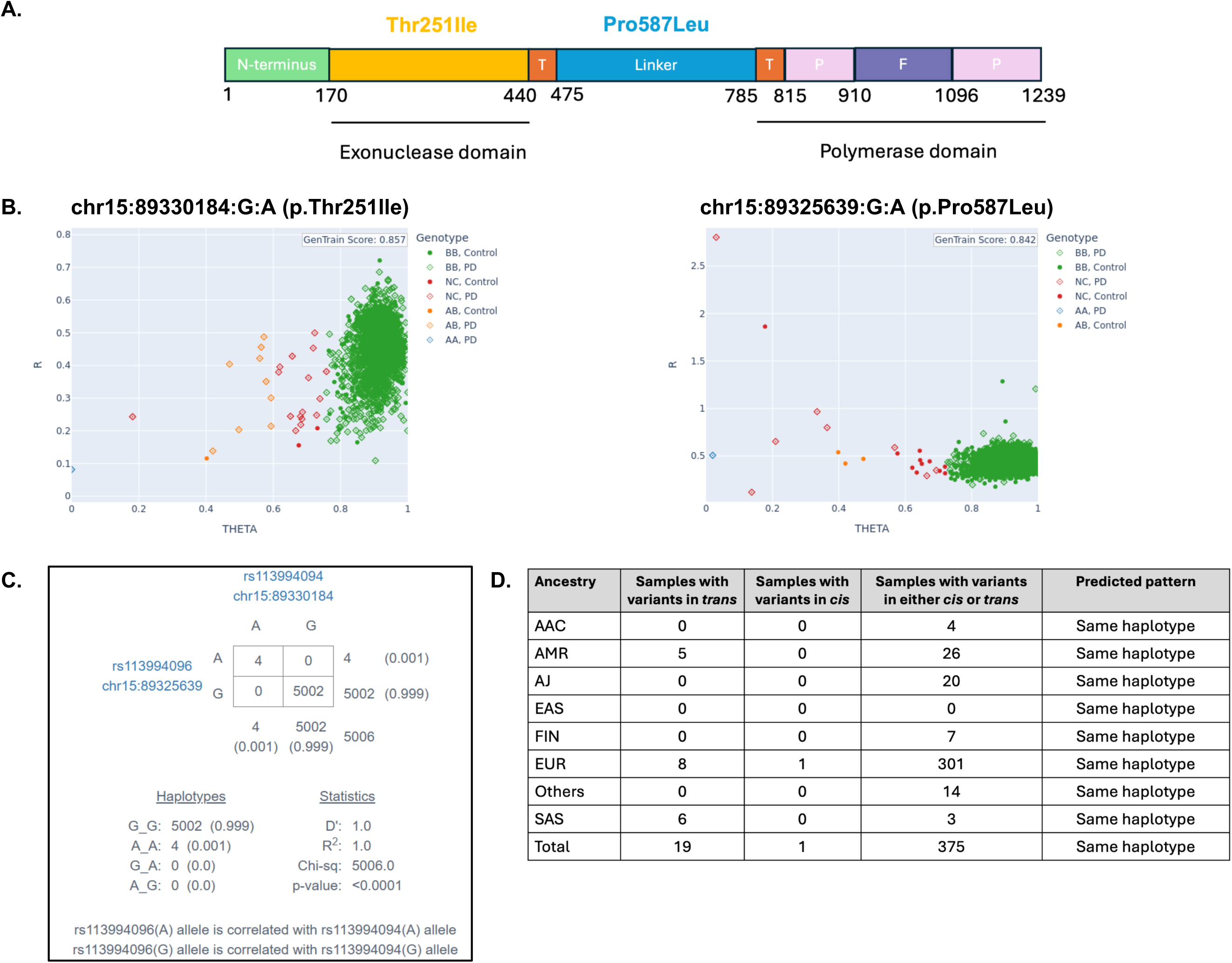
Co-inherited *POLG* variants p.Thr251Ile and p.Pro587Leu. (A) The two co-inherited *POLG* variants [p.Thr251Ile (chr15:89330184:G:A; rs113994094) and p.Pro587Leu (chr15:89325639:G:A; rs113994096)] were mapped onto the domains of the POLG protein. (B) To ensure probe quality, cluster plots for the two variants genotyped on the NBA were visualized. (C) Co-occurrence information from the gnomAD database shows that the A alleles of both variants are inherited on the same haplotype. (D) Ancestry-stratified data adapted from gnomAD show a consistent co-occurrence across populations, supporting their co-inheritance. AAC = African Admixed, AJ = Ashkenazi Jewish, AMR = Latinos and Indigenous People of the Americas, EAS = East Asian, EUR = European, Freq = Frequency, FIN = Finnish, NBA = NeuroBooster Array, MDE = Middle Eastern, SAS = South Asian, WGS = Whole genome sequencing.

**Supplementary Figure 2:**
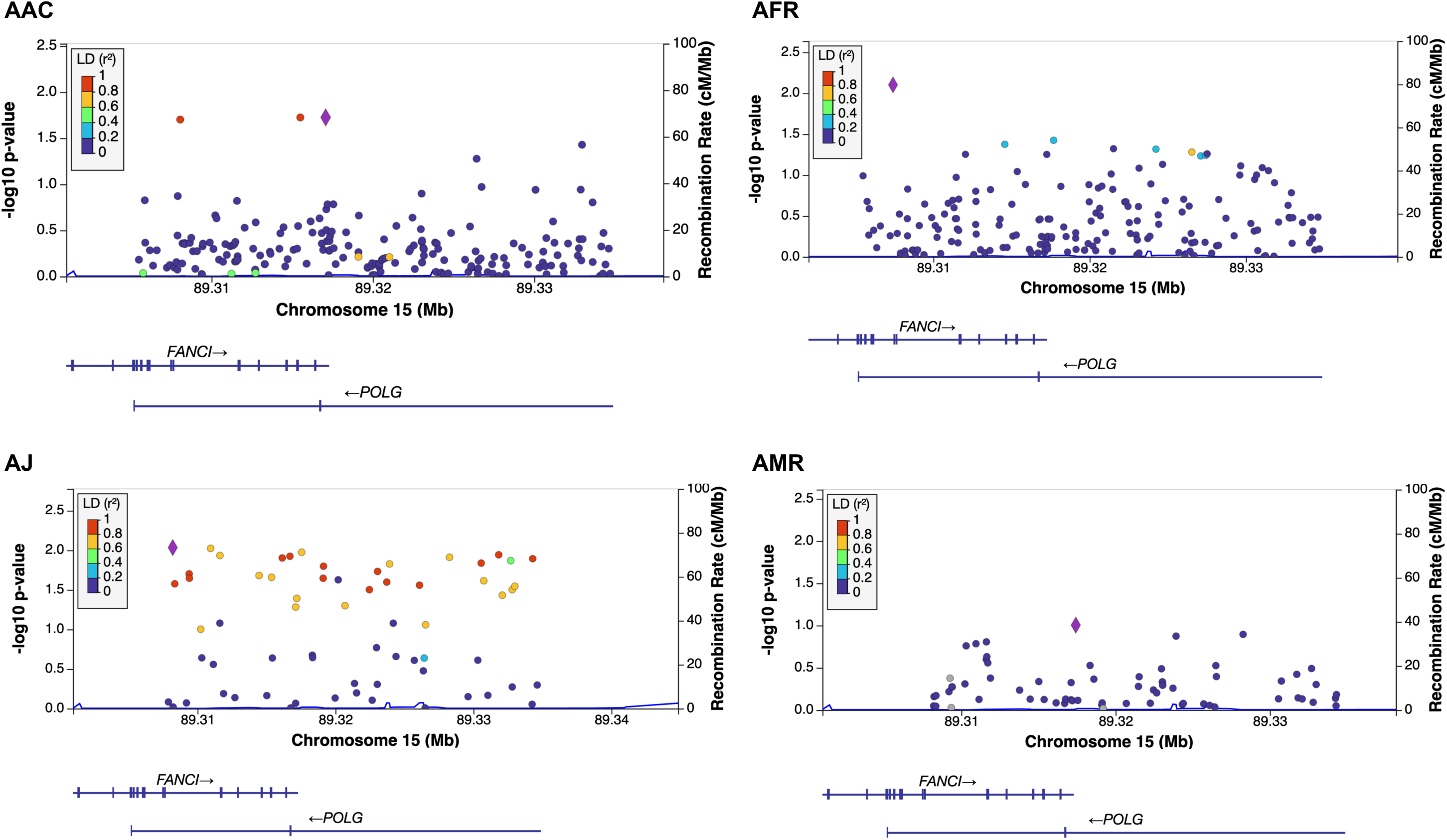
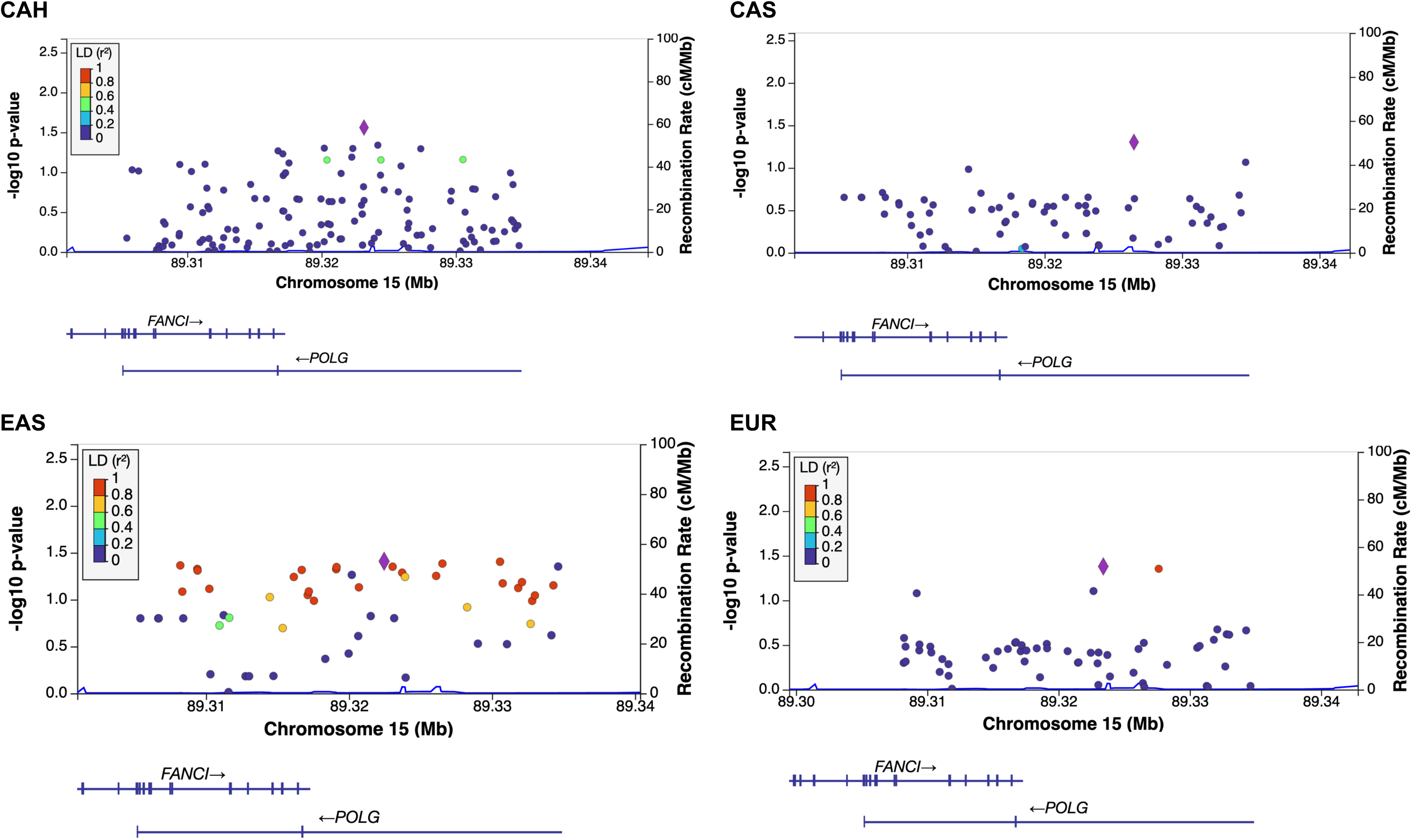
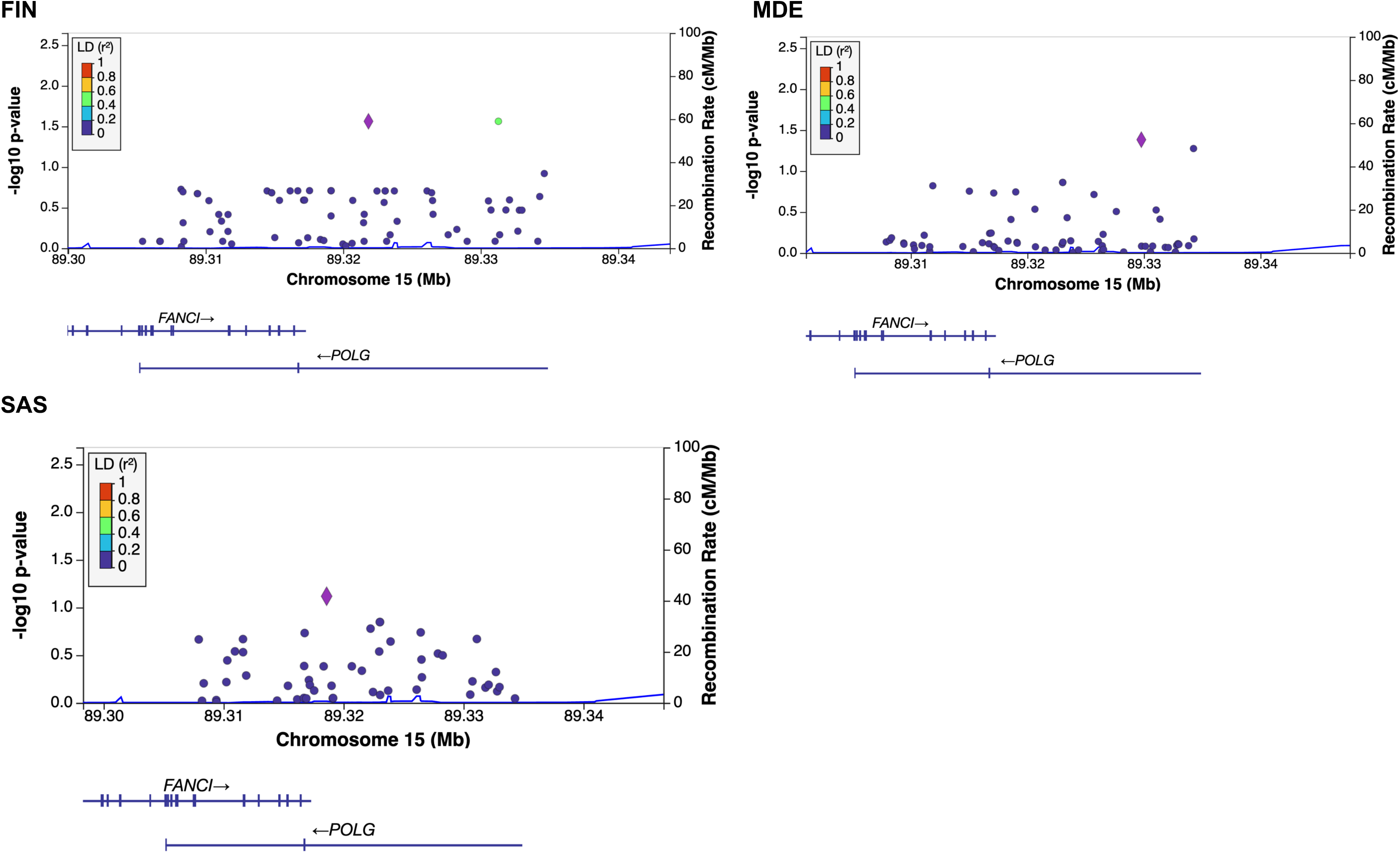
Regional association plot of common *POLG* variants with MAF >1% across ancestries. Multi-ancestry association plots spanning the *POLG* locus are shown. Each point represents the tested common *POLG* variant (MAF >1%) plotted by genomic position (x-axis) and − *log*_10_ (*P*) from case-control logistic regression (y-axis), colored by linkage disequilibrium relative to the lead variant (purple diamond) in each respective cohort using matched 1000 Genomes reference panels. The dashed horizontal line indicates the genome-wide significance threshold (*P* = 5 × 10^−8^), and the blue curve indicates local recombination rate (cM/Mb). AAC = African Admixed, AFR = African, AJ = Ashkenazi Jewish, AMR = Latinos and Indigenous People of the Americas, CAH = Complex Admixture History, CAS = Central Asian, EAS = East Asian, EUR = European, FIN = Finnish, LD = Linkage disequilibrium, MAF = Minor allele frequency, MDE = Middle Eastern, SAS = South Asian.

### Supplementary Tables

**Supplementary Table 1:** Demographics of the studied cohorts from GP2 Release 11 data.

| Genetic ancestry | Data type | Number of subjects, n |  |  |  |
|---|---|---|---|---|---|
|  |  | *N* PD [1] | *N* Other phenotypes [2] | *N* Control [3] | *N* Unaffected [4] |
| AAC | NBA | 491 | 18 | 846 | 3 |
|  | WGS | 190 | 13 | 127 | 2 |
|  | CES | 147 | 0 | 0 | 0 |
| AFR | NBA | 2772 | 44 | 4413 | 1 |
|  | WGS | 1469 | 38 | 2097 | 0 |
|  | CES | 96 | 0 | 0 | 0 |
| AJ | NBA | 2238 | 111 | 804 | 525 |
|  | WGS | 741 | 114 | 162 | 507 |
|  | CES | 800 | 0 | 0 | 0 |
| AMR | NBA | 2229 | 28 | 1508 | 13 |
|  | WGS | 338 | 20 | 170 | 12 |
|  | CES | 320 | 0 | 0 | 0 |
| CAH | NBA | 804 | 33 | 409 | 8 |
|  | WGS | 177 | 50 | 120 | 8 |
|  | CES | 398 | 0 | 0 | 0 |
| CAS | NBA | 1195 | 175 | 1373 | 54 |
|  | WGS | 663 | 165 | 913 | 44 |
|  | CES | 38 | 0 | 0 | 0 |
| EAS | NBA | 4618 | 397 | 2791 | 70 |
|  | WGS | 2697 | 211 | 1049 | 66 |
|  | CES | 94 | 0 | 0 | 0 |
| EUR | NBA | 35955 | 4625 | 25598 | 552 |
|  | WGS | 8044 | 3264 | 1651 | 457 |
|  | CES | 5802 | 0 | 0 | 0 |
| FIN | NBA | 114 | 11 | 20 | 0 |
|  | WGS | 23 | 6 | 4 | 0 |
|  | CES | 16 | 0 | 0 | 0 |
| MDE | NBA | 1101 | 35 | 1309 | 11 |
|  | WGS | 694 | 26 | 976 | 8 |
|  | CES | 42 | 0 | 0 | 0 |
| SAS | NBA | 740 | 103 | 399 | 45 |
|  | WGS | 447 | 67 | 316 | 44 |
|  | CES | 79 | 0 | 0 | 0 |
| **Total** | **NBA** | **52,257** | **5,580** | **39,470** | **1,282** |
|  | **WGS** | **15,483** | **3,974** | **7,585** | **1,148** |
|  | **CES** | **7,832** | **0** | **0** | **0** |

[1] The “PD” category includes individuals with PD from i) unselected PD case/control cohorts, ii) monogenic recruitment, i.e., targeted recruitment of individuals in which a monogenic cause of disease is more likely based on a young age at onset <50 years and/or a positive family history of PD), and iii) genetically enriched cohorts, i.e., targeted recruitment of carriers of pathogenic variants in *GBA1*, *LRRK2*, or *SNCA*.
[2] The “Other phenotypes” category summarizes individuals with phenotypes other than classical PD, including atypical forms of parkinsonism (e.g., multisystem atrophy, progressive supranuclear palsy, corticobasal syndrome, and dementia with lewy bodies) and other neurodegenerative phenotypes (e.g., Alzheimer’s disease, mild cognitive impairment, other forms of dementia).
[3] The “control” category summarizes individuals without PD or other neurological/neurodegenerative phenotypes unrelated to individuals with PD or other phenotypes included in this study.
[4] The “unaffected” category includes unaffected family members of individuals with PD or other phenotypes included in this study.
AAC = African Admixed, AFR = African, AJ = Ashkenazi Jewish, AMR = Latinos and Indigenous People of the Americas, CAH = Complex Admixture History, CAS = Central Asian, CES = Clinical exome sequencing, EAS = East Asian, EUR = European, Freq = Frequency, FIN = Finnish, NBA = NeuroBooster Array, MDE = Middle Eastern, SAS = South Asian, WGS = Whole genome sequencing.

**Supplementary Table 2:** Frequency breakdown of *POLG* CAG repeat alleles in PD patients and controls across ancestries.

| **CAG repeat allele** | **AAC, n (%)** | **AFR, n (%)** | **AJ, n (%)** | **AMR, n (%)** | **CAH, n (%)** | **CAS, n (%)** | **EAS, n (%)** | **EUR, n (%)** | **FIN, n (%)** | **MDE, n (%)** | **SAS, n (%)** |
|---|---|---|---|---|---|---|---|---|---|---|---|
| **PD patients** |  |  |  |  |  |  |  |  |  |  |  |
| **3/10** | 0 (0.00) | 0 (0.00) | 0 (0.00) | 1 (0.13) | 0 (0.00) | 0 (0.00) | 0 (0.00) | 1 (0.01) | 0 (0.00) | 0 (0.00) | 0 (0.00) |
| **4/10** | 0 (0.00) | 0 (0.00) | 0 (0.00) | 0 (0.00) | 0 (0.00) | 0 (0.00) | 0 (0.00) | 1 (0.01) | 0 (0.00) | 0 (0.00) | 0 (0.00) |
| **5/10** | 1 (0.53) | 1 (0.08) | 0 (0.00) | 0 (0.00) | 0 (0.00) | 0 (0.00) | 1 (0.06) | 2 (0.03) | 0 (0.00) | 0 (0.00) | 0 (0.00) |
| **6/10** | 1 (0.53) | 2 (0.15) | 0 (0.00) | 0 (0.00) | 1 (0.54) | 0 (0.00) | 2 (0.13) | 13 (0.17) | 0 (0.00) | 1 (0.12) | 1 (0.35) |
| **7/10** | 0 (0.00) | 1 (0.08) | 1 (0.10) | 10 (1.30) | 2 (1.08) | 0 (0.00) | 17 (1.10) | 32 (0.42) | 0 (0.00) | 1 (0.12) | 3 (1.04) |
| **8/10** | 0 (0.00) | 0 (0.00) | 15 (1.47) | 2 (0.26) | 2 (1.08) | 3 (0.47) | 3 (0.19) | 72 (0.95) | 0 (0.00) | 8 (1.00) | 0 (0.00) |
| **9/10** | 4 (2.11) | 15 (1.15) | 19 (1.86) | 20 (2.59) | 5 (2.69) | 4 (0.63) | 5 (0.32) | 140 (1.85) | 0 (0.00) | 14 (1.74) | 7 (2.42) |
| **4/11** | 0 (0.00) | 0 (0.00) | 0 (0.00) | 0 (0.00) | 0 (0.00) | 0 (0.00) | 0 (0.00) | 0 (0.00) | 0 (0.00) | 1 (0.12) | 0 (0.00) |
| **6/11** | 1 (0.53) | 1 (0.08) | 0 (0.00) | 0 (0.00) | 0 (0.00) | 1 (0.16) | 0 (0.00) | 0 (0.00) | 0 (0.00) | 0 (0.00) | 0 (0.00) |
| **7/11** | 0 (0.00) | 1 (0.08) | 0 (0.00) | 2 (0.26) | 0 (0.00) | 1 (0.16) | 0 (0.00) | 6 (0.08) | 0 (0.00) | 2 (0.25) | 0 (0.00) |
| **8/11** | 0 (0.00) | 0 (0.00) | 3 (0.29) | 1 (0.13) | 0 (0.00) | 0 (0.00) | 0 (0.00) | 7 (0.09) | 0 (0.00) | 0 (0.00) | 0 (0.00) |
| **9/11** | 1 (0.53) | 1 (0.08) | 0 (0.00) | 0 (0.00) | 0 (0.00) | 1 (0.16) | 0 (0.00) | 13 (0.17) | 0 (0.00) | 0 (0.00) | 0 (0.00) |
| **7/12** | 0 (0.00) | 0 (0.00) | 0 (0.00) | 0 (0.00) | 0 (0.00) | 0 (0.00) | 0 (0.00) | 1 (0.01) | 0 (0.00) | 0 (0.00) | 0 (0.00) |
| **8/12** | 0 (0.00) | 0 (0.00) | 0 (0.00) | 0 (0.00) | 0 (0.00) | 0 (0.00) | 0 (0.00) | 1 (0.01) | 0 (0.00) | 0 (0.00) | 0 (0.00) |
| **9/12** | 0 (0.00) | 0 (0.00) | 0 (0.00) | 0 (0.00) | 0 (0.00) | 0 (0.00) | 0 (0.00) | 3 (0.04) | 0 (0.00) | 0 (0.00) | 0 (0.00) |
| **9/13** | 1 (0.53) | 0 (0.00) | 0 (0.00) | 0 (0.00) | 0 (0.00) | 0 (0.00) | 0 (0.00) | 0 (0.00) | 0 (0.00) | 0 (0.00) | 0 (0.00) |
| **7/8** | 0 (0.00) | 0 (0.00) | 0 (0.00) | 0 (0.00) | 0 (0.00) | 0 (0.00) | 0 (0.00) | 0 (0.00) | 0 (0.00) | 1 (0.12) | 0 (0.00) |
| **7/9** | 0 (0.00) | 0 (0.00) | 0 (0.00) | 0 (0.00) | 0 (0.00) | 0 (0.00) | 0 (0.00) | 1 (0.01) | 0 (0.00) | 0 (0.00) | 0 (0.00) |
| **8/9** | 0 (0.00) | 0 (0.00) | 0 (0.00) | 0 (0.00) | 1 (0.54) | 0 (0.00) | 0 (0.00) | 0 (0.00) | 0 (0.00) | 0 (0.00) | 0 (0.00) |
| **9/9** | 0 (0.00) | 0 (0.00) | 0 (0.00) | 1 (0.13) | 0 (0.00) | 0 (0.00) | 0 (0.00) | 1 (0.01) | 0 (0.00) | 0 (0.00) | 0 (0.00) |
| **10/10** | 80 (42.11) | 581 (44.42) | 730 (71.50) | 584 (75.75) | 126 (67.74) | 552 (86.52) | 1462 (94.57) | 5326 (70.26) | 24 (92.31) | 589 (73.35) | 219 (75.78) |
| **10/11** | 58 (30.53) | 381 (29.13) | 211 (20.67) | 101 (13.10) | 30 (16.13) | 61 (9.56) | 52 (3.36) | 1437 (18.96) | 2 (7.69) | 149 (18.56) | 51 (17.65) |
| **10/12** | 28 (14.74) | 154 (11.77) | 21 (2.06) | 31 (4.02) | 12 (6.45) | 13 (2.04) | 1 (0.06) | 342 (4.51) | 0 (0.00) | 26 (3.24) | 5 (1.73) |
| **10/13** | 3 (1.58) | 26 (1.99) | 1 (0.10) | 0 (0.00) | 2 (1.08) | 0 (0.00) | 2 (0.13) | 15 (0.20) | 0 (0.00) | 2 (0.25) | 0 (0.00) |
| **10/14** | 0 (0.00) | 5 (0.38) | 0 (0.00) | 1 (0.13) | 0 (0.00) | 0 (0.00) | 0 (0.00) | 4 (0.05) | 0 (0.00) | 0 (0.00) | 0 (0.00) |
| **10/15** | 0 (0.00) | 0 (0.00) | 0 (0.00) | 0 (0.00) | 0 (0.00) | 0 (0.00) | 0 (0.00) | 1 (0.01) | 0 (0.00) | 0 (0.00) | 0 (0.00) |
| **10/16** | 0 (0.00) | 0 (0.00) | 0 (0.00) | 0 (0.00) | 0 (0.00) | 0 (0.00) | 0 (0.00) | 1 (0.01) | 0 (0.00) | 0 (0.00) | 0 (0.00) |
| **11/11** | 5 (2.63) | 69 (5.28) | 17 (1.67) | 10 (1.30) | 4 (2.15) | 1 (0.16) | 0 (0.00) | 105 (1.39) | 0 (0.00) | 8 (1.00) | 2 (0.69) |
| **11/12** | 5 (2.63) | 53 (4.05) | 2 (0.20) | 6 (0.78) | 1 (0.54) | 1 (0.16) | 1 (0.06) | 48 (0.63) | 0 (0.00) | 1 (0.12) | 1 (0.35) |
| **11/13** | 0 (0.00) | 7 (0.54) | 0 (0.00) | 0 (0.00) | 0 (0.00) | 0 (0.00) | 0 (0.00) | 0 (0.00) | 0 (0.00) | 0 (0.00) | 0 (0.00) |
| **11/14** | 1 (0.53) | 1 (0.08) | 0 (0.00) | 0 (0.00) | 0 (0.00) | 0 (0.00) | 0 (0.00) | 0 (0.00) | 0 (0.00) | 0 (0.00) | 0 (0.00) |
| **12/12** | 0 (0.00) | 8 (0.61) | 1 (0.10) | 1 (0.13) | 0 (0.00) | 0 (0.00) | 0 (0.00) | 7 (0.09) | 0 (0.00) | 0 (0.00) | 0 (0.00) |
| **12/13** | 0 (0.00) | 1 (0.08) | 0 (0.00) | 0 (0.00) | 0 (0.00) | 0 (0.00) | 0 (0.00) | 0 (0.00) | 0 (0.00) | 0 (0.00) | 0 (0.00) |
| **12/14** | 1 (0.53) | 0 (0.00) | 0 (0.00) | 0 (0.00) | 0 (0.00) | 0 (0.00) | 0 (0.00) | 0 (0.00) | 0 (0.00) | 0 (0.00) | 0 (0.00) |
| **Controls** |  |  |  |  |  |  |  |  |  |  |  |
| **5/11** | 0 (0.00) | 1 (0.05) | 0 (0.00) | 0 (0.00) | 0 (0.00) | 0 (0.00) | 0 (0.00) | 0 (0.00) | 0 (0.00) | 0 (0.00) | 0 (0.00) |
| **6/11** | 0 (0.00) | 3 (0.15) | 0 (0.00) | 0 (0.00) | 0 (0.00) | 0 (0.00) | 0 (0.00) | 0 (0.00) | 0 (0.00) | 0 (0.00) | 0 (0.00) |
| **7/11** | 0 (0.00) | 0 (0.00) | 0 (0.00) | 1 (0.33) | 0 (0.00) | 0 (0.00) | 0 (0.00) | 1 (0.08) | 0 (0.00) | 0 (0.00) | 0 (0.00) |
| **8/11** | 0 (0.00) | 0 (0.00) | 0 (0.00) | 1 (0.33) | 0 (0.00) | 0 (0.00) | 0 (0.00) | 0 (0.00) | 0 (0.00) | 0 (0.00) | 0 (0.00) |
| **9/11** | 1 (0.66) | 0 (0.00) | 0 (0.00) | 0 (0.00) | 0 (0.00) | 0 (0.00) | 0 (0.00) | 2 (0.16) | 0 (0.00) | 3 (0.31) | 1 (0.29) |
| **6/13** | 0 (0.00) | 1 (0.05) | 0 (0.00) | 0 (0.00) | 0 (0.00) | 0 (0.00) | 0 (0.00) | 0 (0.00) | 0 (0.00) | 0 (0.00) | 0 (0.00) |
| **3/10** | 0 (0.00) | 1 (0.05) | 0 (0.00) | 0 (0.00) | 0 (0.00) | 2 (0.21) | 0 (0.00) | 1 (0.08) | 0 (0.00) | 0 (0.00) | 0 (0.00) |
| **4/10** | 0 (0.00) | 0 (0.00) | 0 (0.00) | 0 (0.00) | 0 (0.00) | 0 (0.00) | 0 (0.00) | 0 (0.00) | 0 (0.00) | 1 (0.10) | 0 (0.00) |
| **5/10** | 0 (0.00) | 0 (0.00) | 0 (0.00) | 1 (0.33) | 0 (0.00) | 0 (0.00) | 0 (0.00) | 0 (0.00) | 0 (0.00) | 0 (0.00) | 0 (0.00) |
| **6/10** | 1 (0.66) | 3 (0.15) | 0 (0.00) | 0 (0.00) | 0 (0.00) | 1 (0.10) | 2 (0.19) | 0 (0.00) | 0 (0.00) | 1 (0.10) | 4 (1.17) |
| **7/7** | 0 (0.00) | 0 (0.00) | 0 (0.00) | 1 (0.33) | 0 (0.00) | 0 (0.00) | 0 (0.00) | 1 (0.08) | 0 (0.00) | 0 (0.00) | 0 (0.00) |
| **7/10** | 0 (0.00) | 1 (0.05) | 1 (0.39) | 16 (5.23) | 1 (0.81) | 1 (0.10) | 12 (1.16) | 6 (0.49) | 0 (0.00) | 2 (0.21) | 3 (0.87) |
| **8/10** | 1 (0.66) | 4 (0.20) | 4 (1.57) | 2 (0.65) | 2 (1.63) | 1 (0.10) | 0 (0.00) | 11 (0.89) | 0 (0.00) | 9 (0.93) | 1 (0.29) |
| **9/8** | 0 (0.00) | 0 (0.00) | 0 (0.00) | 0 (0.00) | 0 (0.00) | 0 (0.00) | 0 (0.00) | 1 (0.08) | 0 (0.00) | 0 (0.00) | 0 (0.00) |
| **9/10** | 4 (2.65) | 13 (0.65) | 2 (0.78) | 1 (0.33) | 2 (1.63) | 11 (1.13) | 7 (0.68) | 19 (1.54) | 0 (0.00) | 12 (1.24) | 5 (1.46) |
| **10/10** | 75 (49.67) | 890 (44.66) | 193 (75.69) | 247 (80.72) | 84 (68.29) | 825 (84.88) | 951 (92.06) | 893 (72.48) | 4 (66.67) | 682 (70.67) | 245 (71.43) |
| **10/11** | 42 (27.81) | 586 (29.40) | 48 (18.82) | 25 (8.17) | 19 (15.45) | 101 (10.39) | 55 (5.32) | 215 (17.45) | 2 (33.33) | 198 (20.52) | 71 (20.70) |
| **10/12** | 19 (12.58) | 211 (10.59) | 3 (1.18) | 9 (2.94) | 10 (8.13) | 24 (2.47) | 3 (0.29) | 47 (3.81) | 0 (0.00) | 28 (2.90) | 6 (1.75) |
| **10/13** | 2 (1.32) | 49 (2.46) | 0 (0.00) | 0 (0.00) | 0 (0.00) | 0 (0.00) | 1 (0.10) | 1 (0.08) | 0 (0.00) | 0 (0.00) | 2 (0.58) |
| **10/14** | 0 (0.00) | 4 (0.20) | 0 (0.00) | 0 (0.00) | 0 (0.00) | 0 (0.00) | 0 (0.00) | 0 (0.00) | 0 (0.00) | 0 (0.00) | 0 (0.00) |
| **10/15** | 0 (0.00) | 0 (0.00) | 0 (0.00) | 0 (0.00) | 0 (0.00) | 0 (0.00) | 0 (0.00) | 1 (0.08) | 0 (0.00) | 2 (0.21) | 0 (0.00) |
| **11/11** | 5 (3.31) | 101 (5.07) | 4 (1.57) | 1 (0.33) | 0 (0.00) | 5 (0.51) | 2 (0.19) | 19 (1.54) | 0 (0.00) | 22 (2.28) | 4 (1.17) |
| **11/12** | 1 (0.66) | 85 (4.26) | 0 (0.00) | 0 (0.00) | 3 (2.44) | 1 (0.10) | 0 (0.00) | 10 (0.81) | 0 (0.00) | 4 (0.41) | 1 (0.29) |
| **11/13** | 0 (0.00) | 14 (0.70) | 0 (0.00) | 0 (0.00) | 0 (0.00) | 0 (0.00) | 0 (0.00) | 0 (0.00) | 0 (0.00) | 0 (0.00) | 0 (0.00) |
| **11/14** | 0 (0.00) | 3 (0.15) | 0 (0.00) | 0 (0.00) | 0 (0.00) | 0 (0.00) | 0 (0.00) | 0 (0.00) | 0 (0.00) | 0 (0.00) | 0 (0.00) |
| **11/15** | 0 (0.00) | 0 (0.00) | 0 (0.00) | 0 (0.00) | 0 (0.00) | 0 (0.00) | 0 (0.00) | 1 (0.08) | 0 (0.00) | 0 (0.00) | 0 (0.00) |
| **12/12** | 0 (0.00) | 18 (0.90) | 0 (0.00) | 1 (0.33) | 1 (0.81) | 0 (0.00) | 0 (0.00) | 2 (0.16) | 0 (0.00) | 1 (0.10) | 0 (0.00) |
| **12/13** | 0 (0.00) | 5 (0.25) | 0 (0.00) | 0 (0.00) | 1 (0.81) | 0 (0.00) | 0 (0.00) | 1 (0.08) | 0 (0.00) | 0 (0.00) | 0 (0.00) |

**Supplementary Table 3:** Gene-based rare association analyses of *POLG* variants across ancestries.

| **Ancestry** | **N** |  |  | **MAF <1%** |  |  |  |  |  |  |  |  |  |  |  |
|---|---|---|---|---|---|---|---|---|---|---|---|---|---|---|---|
|  |  |  |  | **Potentially functional variants** |  |  | **Coding variants** |  |  | **Loss-of-function variants** |  |  | **Variants with CADD >20** |  |  |
|  | **PD cases** | **Controls** | **Total** | **SKATO** | **CMC-Wald** | **N variant** | **SKATO** | **CMC-Wald** | **N variant** | **SKATO** | **CMC-Wald** | **N variant** | **SKATO** | **CMC-Wald** | **N variant** |
| AFR | 1,344 | 2,049 | 3,393 | 0.0709 | 0.6642 | 141 | 0.1148 | 0.2835 | 85 | 0.4414 | 0.5825 | 3 | 0.2162 | 0.6217 | 57 |
| AJ | 1,023 | 176 | 1,199 | 1.0000 | 0.8143 | 37 | 1.0000 | 0.7130 | 25 | 0.5804 | 0.5335 | 2 | 0.5533 | 0.7667 | 13 |
| AMR | 777 | 296 | 1,073 | 0.7339 | 0.8469 | 80 | 0.6416 | 0.5996 | 41 | 0.8961 | 0.8996 | 1 | 0.7619 | 0.9931 | 25 |
| EAS | 2,163 | 981 | 3,144 | 0.3170 | 0.3298 | 155 | 0.4965 | 0.5350 | 86 | NA | 0.9032 | 2 | 0.4035 | 0.8989 | 61 |
| EUR | 11,569 | 2,040 | 13,609 | 0.8849 | 0.4271 | 320 | 0.8964 | 0.9165 | 195 | 0.3140 | 0.8862 | 12 | 0.8605 | 0.8648 | 131 |
| MDE | 657 | 409 | 1,066 | 0.2697 | 0.2793 | 78 | 0.2077 | 0.3991 | 47 | 0.4533 | 0.8864 | 2 | 0.1514 | 0.3156 | 27 |

Gene-based burden analyses were performed using GP2 Release 11 whole genome sequencing data to evaluate the cumulative contribution of *POLG* variants to PD risk. The number of variants included in the analyses is reported for each ancestry group. P values that reached multiple correction thresholds were highlighted in bold.
AFR = African, AJ = Ashkenazi Jewish, AMR = Latinos and Indigenous People of the Americas, CADD = Combined Annotation Dependent Depletion, EAS = East Asian, EUR = European, Freq = Frequency, MAF = Minor allele frequency, MDE = Middle Eastern, N = Number, NA = Not available, SKAT = Sequence kernel association test, SKATO = Sequence kernel association test - optimal.

**Supplementary Table 4:** Association analysis of common *POLG* exonic variants using GP2 Release 11 genotyping imputed data.

| Variant | GP2 Ancestry | A1 | A1 freq | OR (95% CI) | P-value<br>(BONF) |
|---|---|---|---|---|---|
| chr15:89316763:C:A<br>(p.Gln1236His) | AJ | A | 0.0447 | 0.9271 (0.5880-1.4617) | 0.7444 (1.000) |
|  | AMR |  | 0.0285 | 0.9956 (0.7002-1.4156) | 0.9804 (1.000) |
|  | CAS |  | 0.0309 | 1.1307 (0.7108-1.7986) | 0.6040 (1.000) |
|  | EUR |  | 0.0798 | 1.0331 (0.9722-1.0979) | 0.2939 (1.000) |
|  | MDE |  | 0.0741 | 1.0986 (0.7896-1.5286) | 0.5767 (1.000) |
|  | SAS |  | 0.0117 | 2.3475 (0.6639-8.3008) | 0.1854 (1.000) |
| chr15:89317422:G:T<br>(p.Thr1199=) | AJ | T | 0.0137 | 0.5456 (0.2656-1.1209) | 0.0991 (1.000) |
|  | EUR |  | 0.0106 | 1.0600 (0.9002-1.2481) | 0.4847 (1.000) |
| chr15:89317458:C:G<br>(p.Arg1187=) | AAC | G | 0.0174 | 1.3915 (0.7005-2.7643) | 0.3454 (1.000) |
|  | AFR |  | 0.0319 | 0.8986 (0.7259-1.1122) | 0.3257 (1.000) |
| chr15:89318595:T:C<br>(p.Glu1143Gly) | AAC | C | 0.0115 | 1.0699 (0.4615-2.4805) | 0.8748 (1.000) |
|  | AJ |  | 0.0543 | 0.8528 (0.5732-1.2686) | 0.4319 (1.000) |
|  | CAS |  | 0.0260 | 0.9531 (0.5856-1.5512) | 0.8466 (1.000) |
|  | EUR |  | 0.0414 | 0.9856 (0.9078-1.0699) | 0.7284 (1.000) |
|  | MDE |  | 0.0325 | 0.8082 (0.4973-1.3134) | 0.3900 (1.000) |
|  | SAS |  | 0.0151 | 2.5681 (0.9050-7.2875) | 0.0763 (1.000) |
| chr15:89319006:C:T<br>(p.Thr1066=) | MDE | T | 0.0121 | 0.5753 (0.2564-1.2908) | 0.1800 (1.000) |
|  | SAS |  | 0.0135 | 0.8009 (0.2961-2.1660) | 0.6618 (1.000) |
| chr15:89320789:G:A<br>(p.Tyr986=) | AAC | A | 0.0781 | 1.2915 (0.9221-1.8087) | 0.1367 (1.000) |
|  | AFR |  | 0.1069 | 0.9708 (0.8602-1.0956) | 0.6312 (1.000) |
| chr15:89323415:G:A<br>(p.Leu752=) | EUR | A | 0.0115 | 1.1808 (1.0062-1.3856) | **0.0418** (1.000) |
|  | MDE |  | 0.0145 | 1.3753 (0.6841-2.7645) | 0.3711 (1.000) |
| chr15:89323863:G:T<br>(p.Ala703=) | AAC | T | 0.0313 | 0.8464 (0.4831-1.4827) | 0.5598 (1.000) |
|  | AFR |  | 0.0414 | 1.0800 (0.8925-1.3068) | 0.4290 (1.000) |
| chr15:89324193:C:T<br>(p.Glu662Lys) | AMR | T | 0.0876 | 1.2189 (0.9741-1.5253) | 0.0835 (1.000) |
| chr15:89326688:G:A<br>(p.Arg546Cys) | AAC | A | 0.0168 | 0.9841 (0.4940-1.9605) | 0.9636 (1.000) |
|  | AFR |  | 0.0203 | 0.9364 (0.7126-1.2306) | 0.6374 (1.000) |
| chr15:89328729:G:A<br>(p.Leu376=) | AAC | A | 0.0110 | 0.6223 (0.2361-1.6406) | 0.3376 (1.000) |
|  | AFR |  | 0.0156 | 1.1223 (0.8380-1.5030) | 0.4388 (1.000) |
| chr15:89329018:C:T<br>(p.Lys316=) | CAS | T | 0.0250 | 1.1062 (0.6696-1.8275) | 0.6935 (1.000) |
|  | EAS |  | 0.0111 | 0.7910 (0.5099-1.2273) | 0.2955 (1.000) |
| chr15:89330084:G:A<br>(p.Ile284=) | AFR | A | 0.0167 | 1.2636 (0.9446-1.6904) | 0.1150 (1.000) |
| chr15:89330133:C:G<br>(p.Gly268Ala) | MDE | G | 0.0139 | 0.9213 (0.4371-1.9419) | 0.8295 (1.000) |
| chr15:89333621:T:C<br>(p.Gln45Arg) | AFR | C | 0.0149 | 1.2505 (0.9172-1.7047) | 0.1575 (1.000) |
| chr15:89333627:T:C<br>(p.Gln43Arg) | AAC | C | 0.0409 | 0.8076 (0.4999-1.3046) | 0.3825 (1.000) |
|  | AFR |  | 0.0554 | 1.0339 (0.8735-1.2238) | 0.6986 (1.000) |

A1 = Effect allele, AAC = African Admixed, AFR = African, AJ = Ashkenazi Jewish, AMR = Latinos and Indigenous People of the Americas, CAH = Complex Admixture History, CAS = Central Asian, CES = Clinical exome sequencing, CI = Confidence interval, EAS = East Asian, EUR = European, Freq = Frequency, MDE = Middle Eastern, OR = Odds ratio, SAS = South Asian.

## Notes

### Author Declarations

Data used in the preparation of this manuscript were obtained from the Global Parkinson's Genetics Program (GP2) database accessed via the Terra platform (https://app.terra.bio/#workspaces). For up-to-date information on GP2 data acquisition, access, and policies, visit https://gp2.org/. This project was supported by the Global Parkinson's Genetics Program (GP2; https://gp2.org). GP2 is funded by the Aligning Science Across Parkinson's (ASAP) initiative and implemented by The Michael J. Fox Foundation for Parkinson's Research (MJFF). For a complete list of GP2 members see doi.org/10.5281/zenodo.7904831

### Summary of Updates

We added the results for POLG CAG repeat analyses in the methods, results, discussion, tables and figures.

## References

1. Rahman S, Copeland WC. POLG-related disorders and their neurological manifestations. Nat Rev Neurol 2019;15(1):40–52.

2. Chan SS, Copeland WC. DNA polymerase gamma and mitochondrial disease: understanding the consequence of POLG mutations. Biochim Biophys Acta 2009;1787(5):312–9.

3. Luoma P, Melberg A, Rinne JO, et al. Parkinsonism, premature menopause, and mitochondrial DNA polymerase gamma mutations: clinical and molecular genetic study. Lancet 2004;364(9437):875–82.

4. Bender A, Krishnan KJ, Morris CM, et al. High levels of mitochondrial DNA deletions in substantia nigra neurons in aging and Parkinson disease. Nat Genet 2006;38(5):515–7.

5. Kraytsberg Y, Kudryavtseva E, McKee AC, Geula C, Kowall NW, Khrapko K. Mitochondrial DNA deletions are abundant and cause functional impairment in aged human substantia nigra neurons. Nat Genet 2006;38(5):518–20.

6. Tresse E, Marturia-Navarro J, Sew WQG, et al. Mitochondrial DNA damage triggers spread of Parkinson’s disease-like pathology. Mol Psychiatry 2023;28(11):4902–14.

7. Reeve A, Meagher M, Lax N, et al. The impact of pathogenic mitochondrial DNA mutations on substantia nigra neurons. J Neurosci 2013;33(26):10790–801.

8. Davidzon G, Greene P, Mancuso M, et al. Early-onset familial parkinsonism due to POLG mutations. Ann Neurol 2006;59(5):859–62.

9. Mehta SH, Dickson DW, Morgan JC, Singleton AB, Majounie E, Sethi KD. Juvenile onset Parkinsonism with “pure nigral” degeneration and POLG1 mutation. Parkinsonism Relat Disord 2016;30:83–5.

10. Dolhun R, Presant EM, Hedera P. Novel polymerase gamma (POLG1) gene mutation in the linker domain associated with parkinsonism. BMC Neurol 2013;13:92.

11. Delgado-Alvarado M, de la Riva P, Jimenez-Urbieta H, et al. Parkinsonism, cognitive deficit and behavioural disturbance caused by a novel mutation in the polymerase gamma gene. J Neurol Sci 2015;350(1-2):93–7.

12. Ma L, Mao W, Xu E, et al. Novel POLG mutation in a patient with early-onset parkinsonism, progressive external ophthalmoplegia and optic atrophy. Int J Neurosci 2020;130(4):319–21.

13. Hsieh PC, Wang CC, Tsai CL, Yeh YM, Lee YS, Wu YR. POLG R964C and GBA L444P mutations in familial Parkinson’s disease: Case report and literature review. Brain Behav 2019;9(5):e01281.

14. Calejo M, Vilarinho L, Neiva R, et al. Late-onset Levodopa Responsive Parkinsonism Due to Polymerase γ 1 Mutations. Mov Disord Clin Pract 2018;5(6):645–8.

15. Luoma PT, Eerola J, Ahola S, et al. Mitochondrial DNA polymerase gamma variants in idiopathic sporadic Parkinson disease. Neurology 2007;69(11):1152–9.

16. Hudson G, Tiangyou W, Stutt A, et al. No association between common POLG1 variants and sporadic idiopathic Parkinson’s disease. Mov Disord 2009;24(7):1092–4.

17. Anvret A, Westerlund M, Sydow O, et al. Variations of the CAG trinucleotide repeat in DNA polymerase γ (POLG1) is associated with Parkinson’s disease in Sweden. Neurosci Lett 2010;485(2):117–20.

18. Ylonen S, Siitonen A, Nalls MA, et al. Genetic risk factors in Finnish patients with Parkinson’s disease. Parkinsonism Relat Disord 2017;45:39–43.

19. Eerola J, Luoma PT, Peuralinna T, et al. POLG1 polyglutamine tract variants associated with Parkinson’s disease. Neurosci Lett 2010;477(1):1–5.

20. Gui YX, Xu ZP, Lv W, Liu HM, Zhao JJ, Hu XY. Association of mitochondrial DNA polymerase gamma gene POLG1 polymorphisms with parkinsonism in Chinese populations. PLoS One 2012;7(12):e50086.

21. Balafkan N, Tzoulis C, Müller B, et al. Number of CAG repeats in POLG1 and its association with Parkinson disease in the Norwegian population. Mitochondrion 2012;12(6):640–3.

22. Bentley SR, Shan J, Todorovic M, Wood SA, Mellick GD. Rare POLG1 CAG variants do not influence Parkinson’s disease or polymerase gamma function. Mitochondrion 2014;15:65–8.

23. Cook L, Verbrugge J, Schwantes-An TH, et al. Parkinson’s disease variant detection and disclosure: PD GENEration, a North American study. Brain 2024;147(8):2668–79.

24. Global Parkinson’s Genetics P. GP2: The Global Parkinson’s Genetics Program. Mov Disord 2021;36(4):842–51.

25. Lange LM, Avenali M, Ellis M, et al. Elucidating causative gene variants in hereditary Parkinson’s disease in the Global Parkinson’s Genetics Program (GP2). NPJ Parkinson’s Disease 2023;9(1):100.

26. Towns C, Richer M, Jasaityte S, et al. Defining the causes of sporadic Parkinson’s disease in the global Parkinson’s genetics program (GP2). NPJ Parkinsons Dis 2023;9(1):131.

27. Vitale D, Koretsky MJ, Kuznetsov N, et al. GenoTools: an open-source Python package for efficient genotype data quality control and analysis. G3 (Bethesda) 2025;15(1).

28. Wang K, Li M, Hakonarson H. ANNOVAR: functional annotation of genetic variants from high-throughput sequencing data. Nucleic Acids Res 2010;38(16):e164.

29. Landrum MJ, Lee JM, Riley GR, et al. ClinVar: public archive of relationships among sequence variation and human phenotype. Nucleic Acids Res 2014;42(Database issue):D980–5.

30. Genoox. Franklin by Genoox. [Accessed 6 June 2026]. Available from: https://franklin.genoox.com.

31. Kopanos C, Tsiolkas V, Kouris A, et al. VarSome: the human genomic variant search engine. Bioinformatics 2019;35(11):1978–80.

32. Richards S, Aziz N, Bale S, et al. Standards and guidelines for the interpretation of sequence variants: a joint consensus recommendation of the American College of Medical Genetics and Genomics and the Association for Molecular Pathology. Genet Med 2015;17(5):405–24.

33. DeBalsi KL, Longley MJ, Hoff KE, Copeland WC. Synergistic Effects of the in cis T251I and P587L Mitochondrial DNA Polymerase gamma Disease Mutations. J Biol Chem 2017;292(10):4198–209.

34. Dolzhenko E, Deshpande V, Schlesinger F, et al. ExpansionHunter: a sequence-graph-based tool to analyze variation in short tandem repeat regions. Bioinformatics 2019;35(22):4754–6.

35. Zhan X, Hu Y, Li B, Abecasis GR, Liu DJ. RVTESTS: an efficient and comprehensive tool for rare variant association analysis using sequence data. Bioinformatics 2016;32(9):1423–6.

36. Phillips J, Courel S, Rebelo AP, et al. POLG mutations presenting as Charcot-Marie-Tooth disease. J Peripher Nerv Syst 2019;24(2):213–8.

37. Harrower T, Stewart JD, Hudson G, et al. POLG1 mutations manifesting as autosomal recessive axonal Charcot-Marie-Tooth disease. Arch Neurol 2008;65(1):133–6.

38. Qiu J, Kumar KR, Watson E, Ahmad K, Sue CM, Hayes MW. Dystonia Responsive to Dopamine: POLG Mutations Should Be Considered If Sensory Neuropathy Is Present. J Mov Disord 2021;14(2):157–60.

39. Burusnukul P, de los Reyes EC. Phenotypic variations in 3 children with POLG1 mutations. J Child Neurol 2009;24(4):482–6.

40. Van Goethem G, Dermaut B, Löfgren A, Martin JJ, Van Broeckhoven C. Mutation of POLG is associated with progressive external ophthalmoplegia characterized by mtDNA deletions. Nature Genetics 2001;28(3):211–2.

41. Scuderi C, Borgione E, Castello F, et al. The in cis T251I and P587L POLG1 base changes: description of a new family and literature review. Neuromuscul Disord 2015;25(4):333–9.

42. Lamantea E, Zeviani M. Sequence analysis of familial PEO shows additional mutations associated with the 752C-->T and 3527C-->T changes in the POLG1 gene. Ann Neurol 2004;56(3):454–5.

43. Lamantea E, Tiranti V, Bordoni A, et al. Mutations of mitochondrial DNA polymerase gammaA are a frequent cause of autosomal dominant or recessive progressive external ophthalmoplegia. Ann Neurol 2002;52(2):211–9.

44. Rouzier C, Chaussenot A, Serre V, et al. Quantitative multiplex PCR of short fluorescent fragments for the detection of large intragenic POLG rearrangements in a large French cohort. Eur J Hum Genet 2014;22(4):542–50.

45. Van Goethem G, Martin JJ, Dermaut B, et al. Recessive POLG mutations presenting with sensory and ataxic neuropathy in compound heterozygote patients with progressive external ophthalmoplegia. Neuromuscul Disord 2003;13(2):133–42.

46. Zhou F, Chen J, Zhang T, et al. Characterization of Novel POLG Mutations in Mitochondrial Encephalomyopathy: Pathogenic Validation and Comprehensive Genetic Profiling. Brain Behav 2025;15(11):e71045.

47. Chan SS, Longley MJ, Copeland WC. The common A467T mutation in the human mitochondrial DNA polymerase (POLG) compromises catalytic efficiency and interaction with the accessory subunit. J Biol Chem 2005;280(36):31341–6.

48. Ferrari G, Lamantea E, Donati A, et al. Infantile hepatocerebral syndromes associated with mutations in the mitochondrial DNA polymerase-gammaA. Brain 2005;128(Pt 4):723–31.

49. Horvath R, Hudson G, Ferrari G, et al. Phenotypic spectrum associated with mutations of the mitochondrial polymerase gamma gene. Brain 2006;129(Pt 7):1674–84.

50. Azrak S, Ayyasamy V, Zirpoli G, et al. CAG repeat variants in the POLG1 gene encoding mtDNA polymerase-gamma and risk of breast cancer in African-American women. PLoS One 2012;7(1):e29548.

51. Rani DS, Carlus SJ, Poongothai J, et al. CAG repeat variation in the mtDNA polymerase γ is not associated with oligoasthenozoospermia. International Journal of Andrology 2009;32(6):647–55.

52. Aknin-Seifer IE, Touraine RL, Lejeune H, et al. Is the CAG repeat of mitochondrial DNA polymerase gamma (POLG) associated with male infertility? A multi-centre French study. Hum Reprod 2005;20(3):736–40.

